# Comparison of methods for characterizing skin pigment diversity in research cohorts

**DOI:** 10.1101/2025.02.21.25322707

**Authors:** Michael S. Lipnick, Danni Chen, Tyler Law, Kelvin Moore, Jenna C. Lester, Ellis P. Monk, Carolyn M. Hendrickson, Yu Chou, Caroline Hughes, Ella Behnke, Seif Elmankabadi, Lily Ortiz, Fekir Negussie, Gregory Leeb, Odinakachukwu Ehie, Isabella Auchus, Elizabeth N. Igaga, Ronald Bisegerwa, Olubunmi Okunlola, Philip Bickler, John Feiner, Leonid Shmuylovich

## Abstract

**Background:** Some pulse oximeters perform worse in people with darker skin, and this may be due to inadequate diversity of skin pigment in device development study cohorts. Guidance is needed to accurately and equitably characterize skin pigment to ensure diversity in research cohorts. We tested multiple methods for characterizing skin pigment to assess comparability and impact on cohort diversity.

**Objectives:**

- Assess reliability and comparability of common skin pigment measurement methods
- Compare findings from different anatomical sites
- Demonstrate that pigment cannot be assumed from US National Institutes for Health (NIH) race categories

**Methods:** We used three subjective methods (perceived Fitzpatrick pFP, Monk Skin Tone MST and Von Luschan VL) and two objective methods (Konica Minolta CM-700d spectrophotometer and Delfin Skin Color Catch DSCC colorimeter) for individual typology angle (ITA), across multiple measurement sites in adults. We calculated ΔE to estimate operator perceptibility thresholds for subjective methods and to determine reproducibility for objective methods. We used each method to categorize participants as ‘light, medium, or dark’ and compared the impact of method selection on cohort diversity.

**Results:** We studied 789 participants, with 33,856 assessments. The MST had the widest luminosity range, and VL had the least discernible adjacent categories. With ‘dark’ defined as ITA <-30°, 14% of participants were categorized ‘dark’ as compared to 26% by pFP or 16% by MST. Approximately half of the ‘dark’ cohort had an ITA <-50°. With an ITA threshold <-50°, only 7% of the cohort was categorized as ‘dark.’ When ‘Black or African American’ self-identification was used to define ‘dark’, 23% of the cohort was categorized as such. Each self-assigned NIH race category included a wide range of ITA and subjective scale categories. Both ITA and L* from the KM-700d and DSCC demonstrated strong correlation (⍴ > 0.7).

**Conclusion:** Common methods for skin pigment characterization, especially the use of race or subjective scales, have significant limitations. When applied to the same cohort, different methods yield significantly different results, and some may overestimate diversity. Previously published ITA thresholds for defining ‘dark’ skin are too light and lead to underrepresentation of people with darker skin.

## Introduction

Pulse oximeters do not work as well in people with dark skin and recently this bias has been linked to health disparities.^1–4^ In the past, some regulatory agencies explicitly encouraged inclusion of “lightly” and “darkly” pigmented subjects in oximeter verification trials, but never defined these terms or how to measure them.^5^ This ambiguity may have allowed oximeters to reach the market with false reassurance of equitable performance across a diverse range of pigmentation.^6^ There is an urgent need for data-driven guidance on pigmentation assessment to ensure diverse representation in optical medical device verification studies.

Broadly speaking, skin pigmentation can pragmatically be assessed through either subjective methods, such as matching skin tone to color scales, or objective methods, such as spectrophotometry or colorimetry.^7,8^ It is not clear, however, which approach best ensures diversity of pigmentation in a study population or how to communicate this diversity in a manner that is easy to understand. Are subjective methods reliable? Do different subjective and objective tools yield similar results? What cutoffs should be used for objective measures? And, how do anatomic measurement sites and self-reported US National Institutes of Health (NIH) race impact findings?

This study aimed to answer these questions by using multiple objective and subjective methods to assess skin pigmentation in a diverse study cohort.

## Materials & Methods

We prospectively collected data from: 1. A ‘lab cohort’ of healthy adults enrolled for oximeter verification studies at the UCSF Hypoxia Lab; and 2. An ‘ICU cohort’ of critically-ill adults enrolled in the EquiOx Clinical Trial (ClinicalTrials.gov NCT06142019) at San Francisco General Hospital, a public safety net hospital.^12^ Study sites were chosen based on convenience and with the intent to capture the local diversity of pigment. Sample size was not prespecified, and we used all available data from the trial period. The UCSF IRB approved this study (#21-35637, #22-36553 and #23-40212).

Skin pigmentation was assessed with subjective and objective methods. For subjective methods, we chose the Von Luschan (VL), Monk Skin Tone (MST), and perceived Fitzpatrick (pFP) scales. While there is no standard visual Fitzpatrick Scale^9^, we implemented a 6-category ‘perception’ of the scale that is found online.^10^ The VL and pFP were printed with a Canon Pro4000 on tier-1 bright white 330gm paper, and the MST scale was printed by a professional photo printing service. ^11^ For objective methods, we used the Konica Minolta CM-700d spectrophotometer (KM-700d) and the Delfin Skin Color Catch (DSCC) tristimulus colorimeter. The DSCC and VL were only used in the ICU cohort.

We divided each subjective scale into approximate thirds to define ‘light’ (pFP I/II, vL 1-15, MST A-C), ‘medium’ (pFP III/IV, vL 16-29, MST D-G), and ‘dark’ (pFP V/VI, vL 29-36, MST H-J). In the ICU cohort, one research coordinator assessed participants under overhead fluorescent lighting, and in the lab cohort two research coordinators (and a third to resolve discordance) assessed participants under a full spectrum light source (Aputure Amaran 60 XS 6500k).^12^

Race was self-identified or extracted from the medical record, using five NIH categories for race with the addition of a sixth category ‘other.’^13^

The spectrophotometer and colorimeter were calibrated per manufacturers’ recommendations. At each anatomic site, we obtained three measurements after placing the device against the skin with enough pressure to seal the aperture, except at the fingernail where this was not always possible. Anatomical sites included sun-exposed, sun-protected, and commonly used locations for pulse oximeters.

We used KM-700d with 3 mm aperture and CM-SA Skin Analysis Software (Konica Minolta) with illuminant ‘D65’ and observer angle 10° before exporting CIELAB L*, a*, b* color space coordinates. Individual typology angle (ITA) was calculated as the angle in the L*b* plane between (0,50) and (b*, L*):

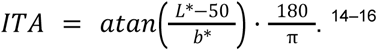

For both devices, we converted L*a*b* to RGB to help visualize L*a*b* colorspace using the MATLAB *lab2rgb*() function, or the Python python-colormath package (3.0.0), with D65 white point and sRGB colorspace. We used previously published ITA categories (very light >55° > light > 41° > intermediate >28° > tan >10° > brown > -30° > dark) to assign light (ITA > 41°), medium (41° > ITA > -30°) and dark (ITA < -30°).^14^

### Statistical Analysis

Agreement between triplicate values was determined by calculating ΔE between each L*a*b* measurement and the median L*a*b* (using the CIE76 euclidean distance formula)^17^. Measurements with ΔE >10 (>10-fold higher than expected ΔE for spectrophotometer repeat measures) or negative b* were excluded.

To estimate discernibility between adjacent subjective scale categories, we converted published or measured RGB values to L*a*b for each scale, then calculated ΔE. We defined ΔE < 2 as indistinguishable.^18^

To compare how different methods impact distribution of lightly and darkly pigmented subjects, histograms were compared visually and numerically by kurtosis. Paired t-tests compared objective measurements between anatomic sites in each subject. Strength of correlation of objective measurements between anatomical sites was evaluated byspearman’s rho (⍴, ⍴ > 0.6 indicates strong correlation). The ΔE and ΔITA were used to assess repeatability and inter-device comparability. Analyses were performed using MATLAB (The Mathworks, Inc., Boston, MA), STATA v 17·0 (Statacorp., College Station, TX), and Python v 3.9.6 (Python Software Foundation, 2024).

## Results

### Demographics

This study included 789 participants and 33,856 skin assessments. In the lab cohort, 57 subjects participated on multiple days (mean 99.91 days (95% CI: 80.9 - 118.92) between assessments). Demographics are presented in Table 1, and a breakdown of measurements by anatomical sites is presented in Table 2.

**Table 1.** Study participant demographics and skin pigment measurement methods.

|  | **ICU Cohort**<br>**(N = 637)** | **Lab Cohort**<br>**(N = 152)** | **All**<br>**(n = 789)** |
| --- | --- | --- | --- |
| **Age (Years)** | 61.0 ± 17.1 | 27.2 ± 5.4 | 54.6 ± 20.4 |
| **Assigned Sex** |  |  |  |
| - Male | 470 (73.8) | 83 (54.6) | 553 (70.1) |
| - Female | 160 (25.1) | 65 (42.8) | 225 (28.5) |
| - Not Reported | 7 (1.1) | 4 (2.6) | 11 (1.4) |
| **Bilirubin** | 1.1 ± 2.2 | NA | 1.1 ± 2.2 |
| **Hgb** | 10.8 ± 2.3 | 13.6 ± 1.6 | 11.2 ± 2.4 |
| **NIH Race** |  |  |  |
| - American Indian or Alaska Native | 6 (0.9) | 0 (0) | 6 (0.8) |
| - Asian | 131 (20.6) | 38 (25.0) | 169 (21.4) |
| - Black or African American | 137 (21.5) | 39 (25.7) | 176 (22.3) |
| - Native Hawaiian or Other Pacific Islander | 11 (1.7) | 0 (0) | 11 (1.4) |
| - Other/Multi | 193 (30.3) | 30 (19.7) | 223 (28.3) |
| - White | 159 (25.0) | 45 (29.6) | 204 (25.9) |
| **Assessed by pFP n(%)** | 632 (99.2) | 118 (77.6) | 750 (95.1) |
| **Assessed by VL n(%)** | 637 (100) | NA | 637 (80.7) |
| **Assessed by MST n(%)** | 516 (81.0) | 125 (82.2) | 641 (81.2) |
| **Assessed by CM-700d n(%)** | 598 (93.9) | 152 (100) | 750 (95.1) |
| **Assessed by DSCC n(%)** | 263 (41.3) | NA | 42 (5.3) |

This table presents descriptive statistics for age, assigned sex, bilirubin, hemoglobin (Hgb), self-reported race (or ethnic origin), and the total number of participants assessed using the five different skin pigment measurement methods. Data are presented separately for the ICU and lab cohorts as well as for the combined cohort. Continuous variables are presented as mean ± SD, and categorical variables as n (%).

**Table 2.**
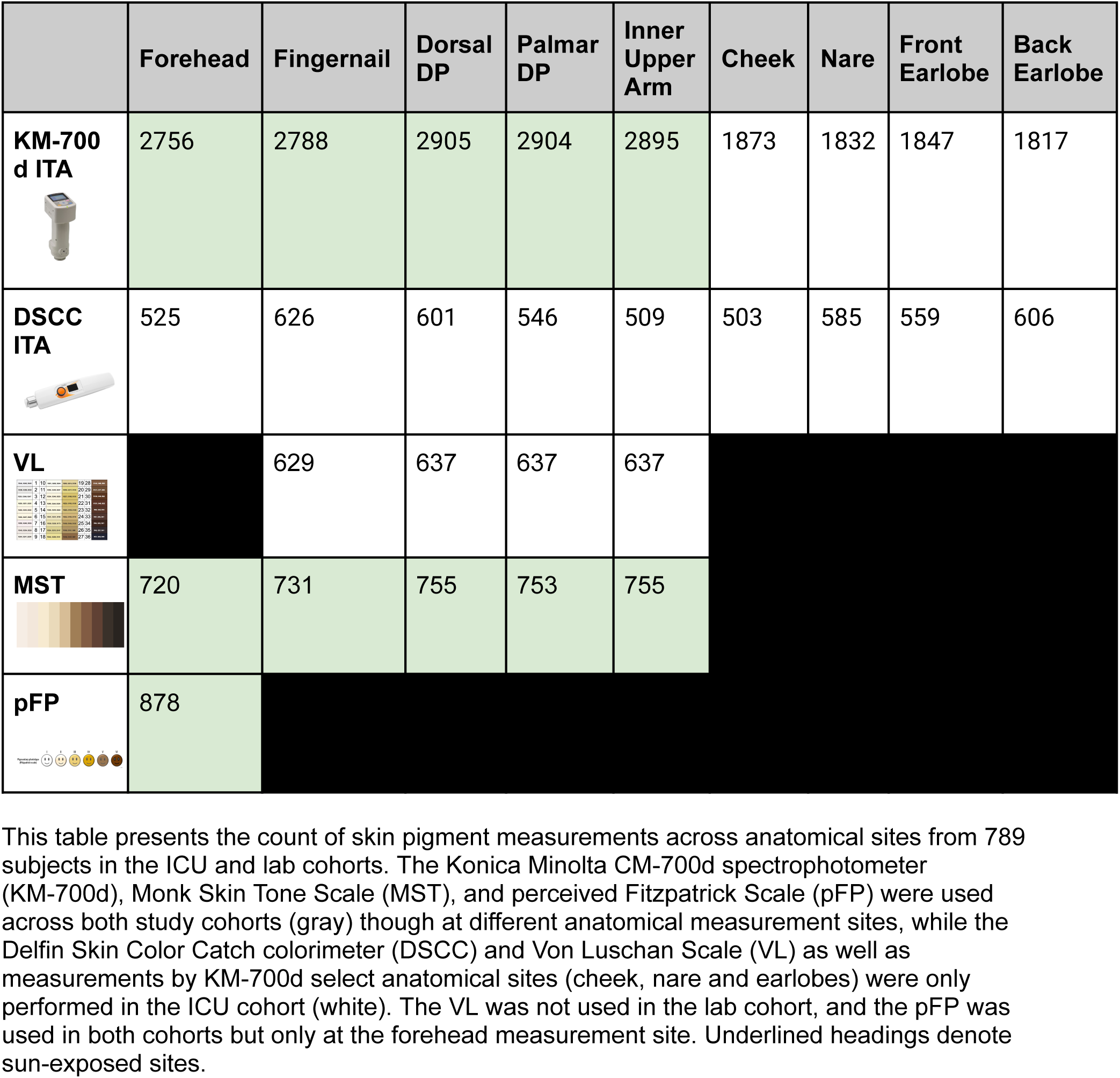
Skin pigment measurement count by anatomical site.

|  | Forehead | Fingernail | Dorsal DP | Palmar DP | Inner Upper Arm | Cheek | Nare | Front Earlobe | Back Earlobe |
| --- | --- | --- | --- | --- | --- | --- | --- | --- | --- |
| **KM-700 d ITA** | 2756 | 2788 | 2905 | 2904 | 2895 | 1873 | 1832 | 1847 | 1817 |
| **DSCC ITA** | 525 | 626 | 601 | 546 | 509 | 503 | 585 | 559 | 606 |
| **VL** |  | 629 | 637 | 637 | 637 |  |  |  |  |
| **MST** | 720 | 731 | 755 | 753 | 755 |  |  |  |  |
| **pFP** | 878 |  |  |  |  |  |  |  |  |

This table presents the count of skin pigment measurements across anatomical sites from 789 subjects in the ICU and lab cohorts. The Konica Minolta CM-700d spectrophotometer (KM-700d), Monk Skin Tone Scale (MST), and perceived Fitzpatrick Scale (pFP) were used across both study cohorts (gray) though at different anatomical measurement sites, while the Delfin Skin Color Catch colorimeter (DSCC) and Von Luschan Scale (VL) as well as measurements by KM-700d select anatomical sites (cheek, nare and earlobes) were only performed in the ICU cohort (white). The VL was not used in the lab cohort, and the pFP was used in both cohorts but only at the forehead measurement site. Underlined headings denote sun-exposed sites.

### Subjective skin pigment measurement methods

#### Comparison of luminosity and perceptibility thresholds

Subjective scales differed in their ranges of light to dark (luminosity = L*, from 0 to 100) and ITA (Figure 1a). The lowest (darkest) L* for MST (MST J=14.7) was less than the lowest L* for pFP (pFP VI = 15.9) or VL (vL 36 = 28.9), and the largest (lightest) L* for MST (MST A = 94.3) was larger than L* for pFP (pFP I = 85.8) or VL (vL 1= 88.6).

**Figure 1a.**
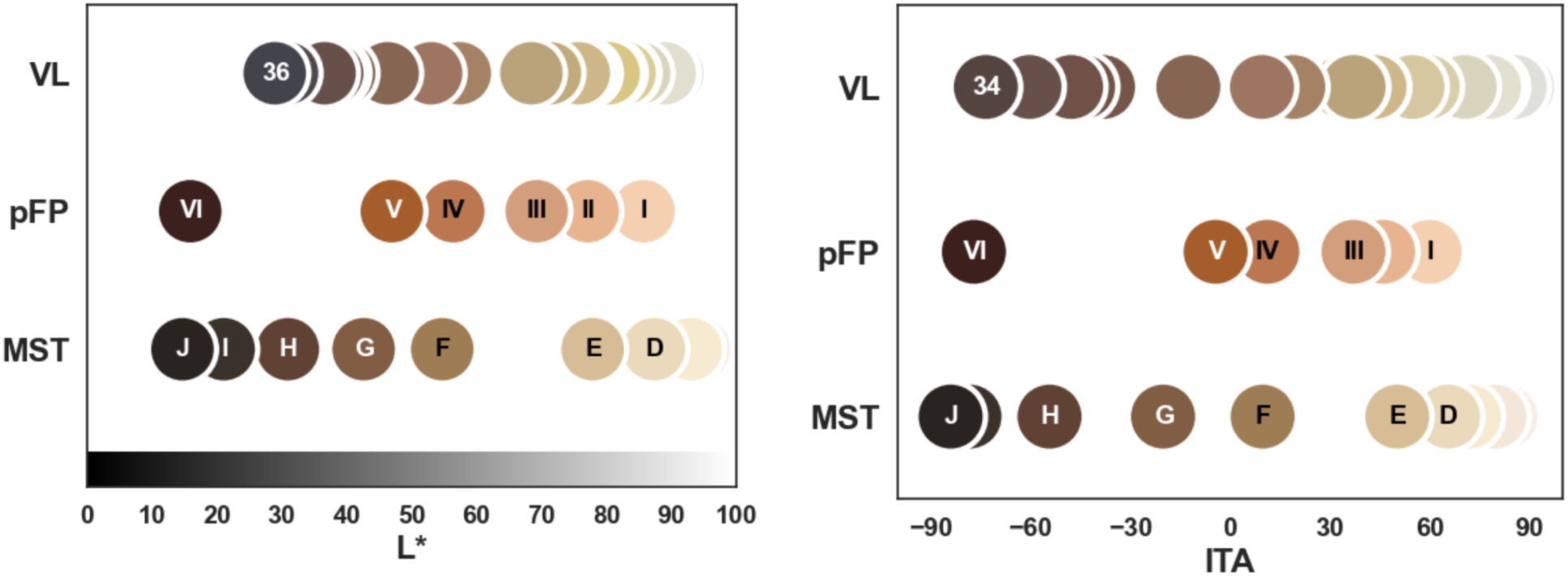
Comparison of subjective scale luminosity and ITA. Luminosity (L*) (*left*) and individual typology angle (ITA) (*right*) derived from the L*a*b* coordinates are compared for each category from the Monk Skin Tone Scale (MST), perceived Fitzpatrick Scale (pFP), and Von Luschan Scale (VL) (*bottom*). The L*a*b* values were obtained from published values from MST, and from measured values for pFP and VL scales. Circles are colored based on published RGB values for MST and measured L*a*b* converted to RGB for VL and PFP. Six VL categories with b*<0 were excluded from this figure.

Subjective scales differed in the distinguishability of adjacent categories. All adjacent categories of pFP and MST scales had ΔE > 2 (mean ΔE_pFP_=19.08±15.64, mean ΔE_MST_=11.26±6.35 (Figure 1b), while the VL scale had 10 categories with ΔE < 2 (i.e., indistinguishable). The MST scale categories at the extremes had lower adjacent ΔE than mean ΔE_MST_(ΔE_A-B_ = 3.02, ΔE_I-J_ = 6.77).

**Figure 1b.**
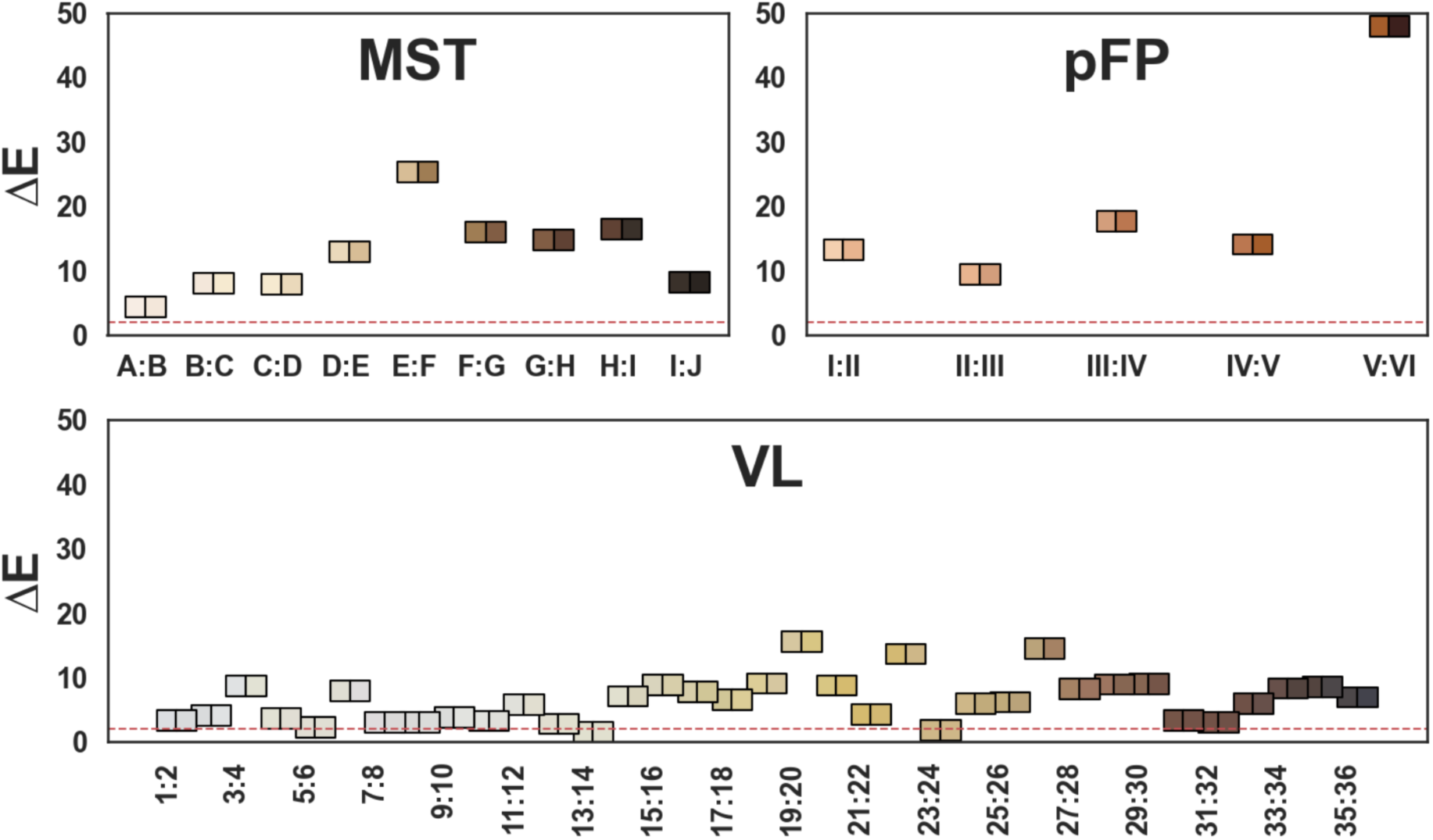
Perceptibility thresholds for subjective scale categories. This figure compares the ΔE values (CIE76) derived from the L*a*b* coordinates of adjacent categories from the Monk Skin Tone Scale (MST) (top left), perceived Fitzpatrick Scale (pFP)(top right), and Von Luschan Scale (VL). The ΔE values are represented by the bottom of each square. The red dashed line represents a ΔE of 2, which is commonly used as a threshold for perceptibility of color differences by a typical observer. Tiles are colored based on published RGB values for MST and measured L*a*b* converted to RGB for VL and PFP.

#### Variability of pigmentation categorization (light, medium, or dark) using MST, pFP, and VL

Among subjects with repeat measurements across time, subjective assessment provided consistent results with median range of both MST and pFP across study days of 1.0 (IQR: 0.0 - 1.0). Categorization of participants as light, medium, or dark was not consistent between scales (Figure 2a). For example, 41% of subjects categorized as dark by pFP (V-VI) were categorized as medium by MST (D-G). Subjects categorized as medium by MST (D-G), spanned the entire range of pFP. When comparing MST or pFP to VL, all scales had many overlapping categories within the same participant (Figure S1 and Figure S2).

**Figure 2.**
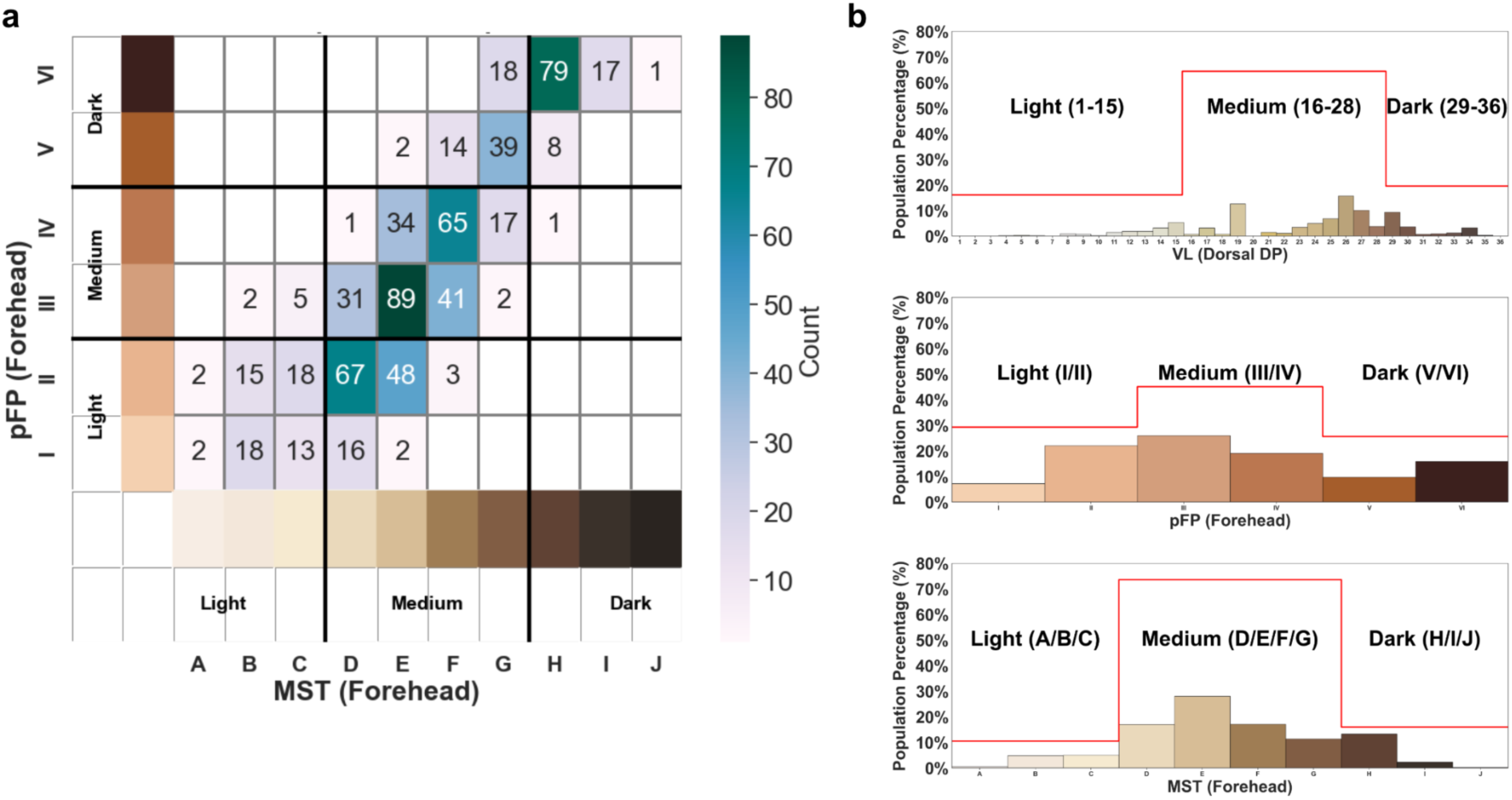
Study cohort diversity characterized by different subjective scales. Panel 2a displays a heatmap to visualize comparison of Monk Skin Tone Scale (MST) at the forehead and perceived Fitzpatrick Scale (pFP) at the forehead of the same participants to demonstrate categorization as light, medium, or dark was not consistent between scales. Panel 2b compares the proportion of the study cohort characterized as light, medium or dark when using different subjective skin pigment assessment methods: MST at the forehead, pFP at the forehead, and Von Luschan Scale (VL) at the dorsal distal phalanx. The x-axes represent the values for each scale, and the y-axes show the population percentage within each category. Red stepped lines in each subplot outline the cumulative proportions of the cohort within the defined subgroups.

The proportion of the study cohort categorized as dark varied by scale with 26% by pFP (ICU and lab cohorts combined), 16% by MST (ICU and lab cohorts combined), and 19% by VL (ICU cohort) (Figure 2b). The proportions also varied depending on cutoffs, for example using pFP VI (rather than V/VI) to define ‘dark’ decreases the dark pigmentation cohort from 26% to 16%. The distribution of subjects across pigmentation categories also varied by scale, with the distribution of pFP being nearly flat (kurtosis -0.93) and the distribution for VL and MST being closer to normal (kurtosis -0.26 and -0.37, respectively)

#### Variability of pigmentation categorization (light, medium, or dark) by assessment site

The distribution of differences in MST measured at different anatomic sites in the same subject (ΔMST) is shown in Figure S3. When comparing forehead and dorsal DP, ΔMST is centered at 0 and the assignment of subjects to light, medium, and dark at these sites is consistent (Figure S4a), indicating that perceived pigmentation at these two sun-exposed sites was in close agreement. In contrast, ΔMST has a positive skew when comparing forehead or dorsal DP to inner upper arm, palmar DP, or fingernail, suggesting that forehead and dorsal DP were consistently judged to be darker by MST. This positive skew in ΔMST impacted the assignment of subjects to light, medium, or dark (Figure S4b).

### Objective skin pigment measurement methods

#### Inter- and intra-device variability

Assessment of triplicate measurements at the forehead for the same participant showed a mean ΔITA of 0.8 ± 1.6 and ΔE 0.6 ± 0.6 for the KM-700d, and ΔITA 1.0 ± 1.8 and ΔE 1.4 ± 1.0 for the DSCC. Sites that were challenging to measure (fingernail, front earlobe, and back earlobe) exhibited higher ΔITA (ΔITA 2.5 ± 5.2, 1.6 ± 3.3, 2.5 ± 4.9, respectively) than the triplicate ΔITA measured at the forehead (1.0 ± 1.8) (Figures S5-S8). Assessment of ITA by the KM-700d and DSCC demonstrated a strong inter-device correlation (⍴ > 0.6) across all sites (Figure 3).

**Figure 3.**
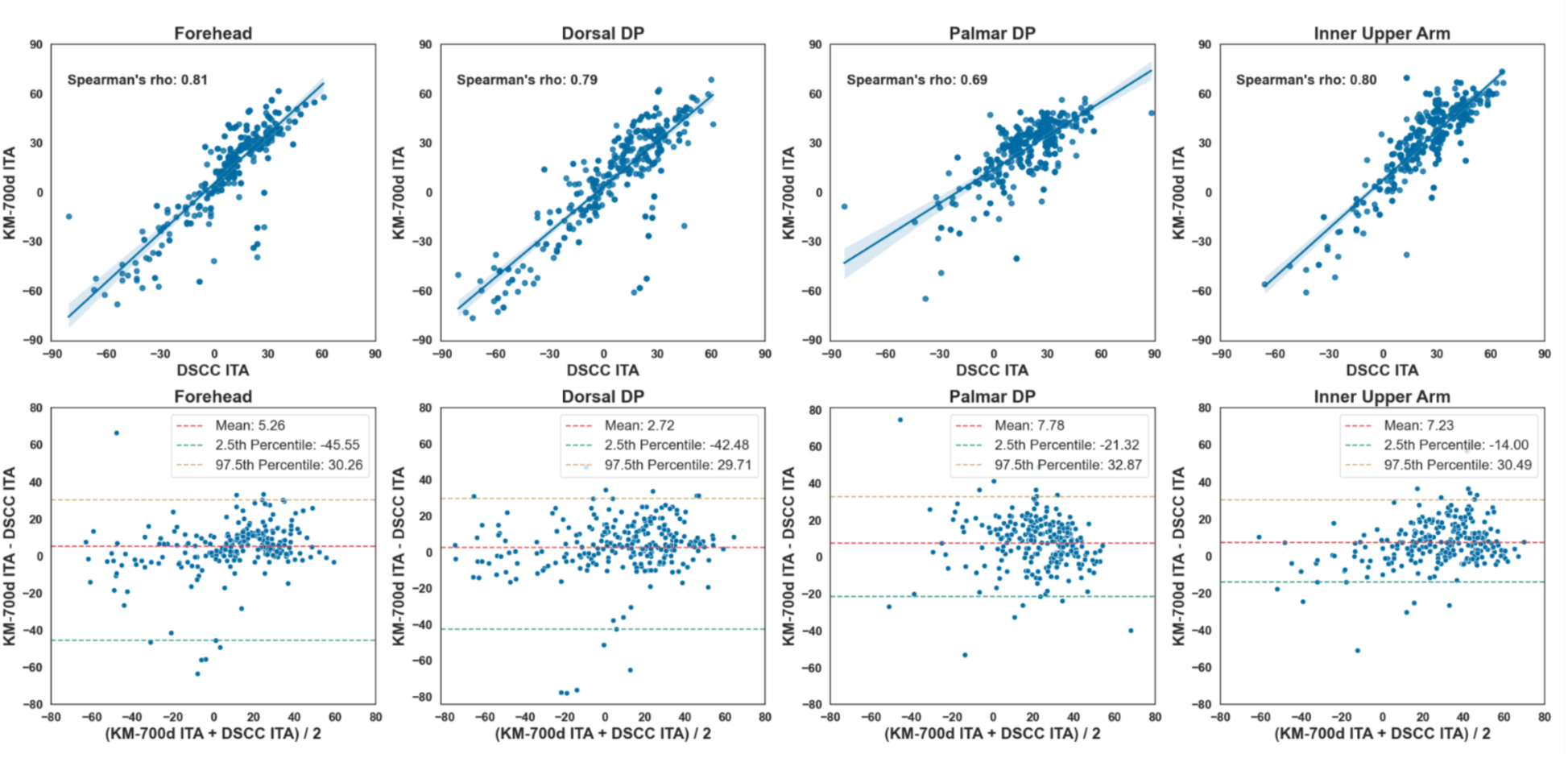
Comparison of individual typology angle between two objective methods across anatomical sites. This figure consists of a four by two grid of subplots, with each column representing an anatomical site: forehead, dorsal distal phalanx (DP), palmar DP, and inner upper arm. The top row displays regression plots, where Delfin Skin Color Catch (DSCC) colorimeter individual typology angle (ITA) is plotted on the x-axis and Konica Minolta CM-700d (KM-700d) ITA on the y-axis. Each regression plot includes a shaded area to visualize the 95% confidence interval for the estimated regression line and an annotation of Spearman’s rho for the correlation between the two devices. The bottom row contains Bland-Altman (BA) plots, with the x-axis representing the mean of DSCC ITA and KM ITA and the y-axis showing the difference between them. The BA plots are annotated with the mean difference and the 2.5th and 97.5th percentiles to illustrate the limits of agreement. Data shown are from ICU cohort only, as the DSCC device was not used in the lab cohort.

Among subjects with repeat skin measurements over time, objective assessment provided consistent measures with a mean range of ITA across study days of 11.9° (95% CI: 10.8° - 12.9°).

#### Variability of pigmentation categorization (light, medium, or dark) by ITA threshold

In Figure 4a, applying previously published ITA categories to forehead ITA leads to a distribution of 13.6% dark, 75.9% medium, and 10.5% light . For the 13.6% categorized as dark (ITA<-30°, n=116), ITA spanned from -30.1° to -75.8°, with 47.4% (n=55) having an ITA<-50°. Across both cohorts, only 0.7% (n=6) of participants were categorized as ‘very light’ (ITA > 55°). Setting alternative thresholds of ITA >30° for ‘light’ and <-50° as ‘dark’ shifts the distribution considerably(Figure 4b). For 262 subjects measured by both KM-700d and DSCC, the distribution of ITA values (at the same anatomic site) were indistinguishable (Figure S9).

**Figure 4.**
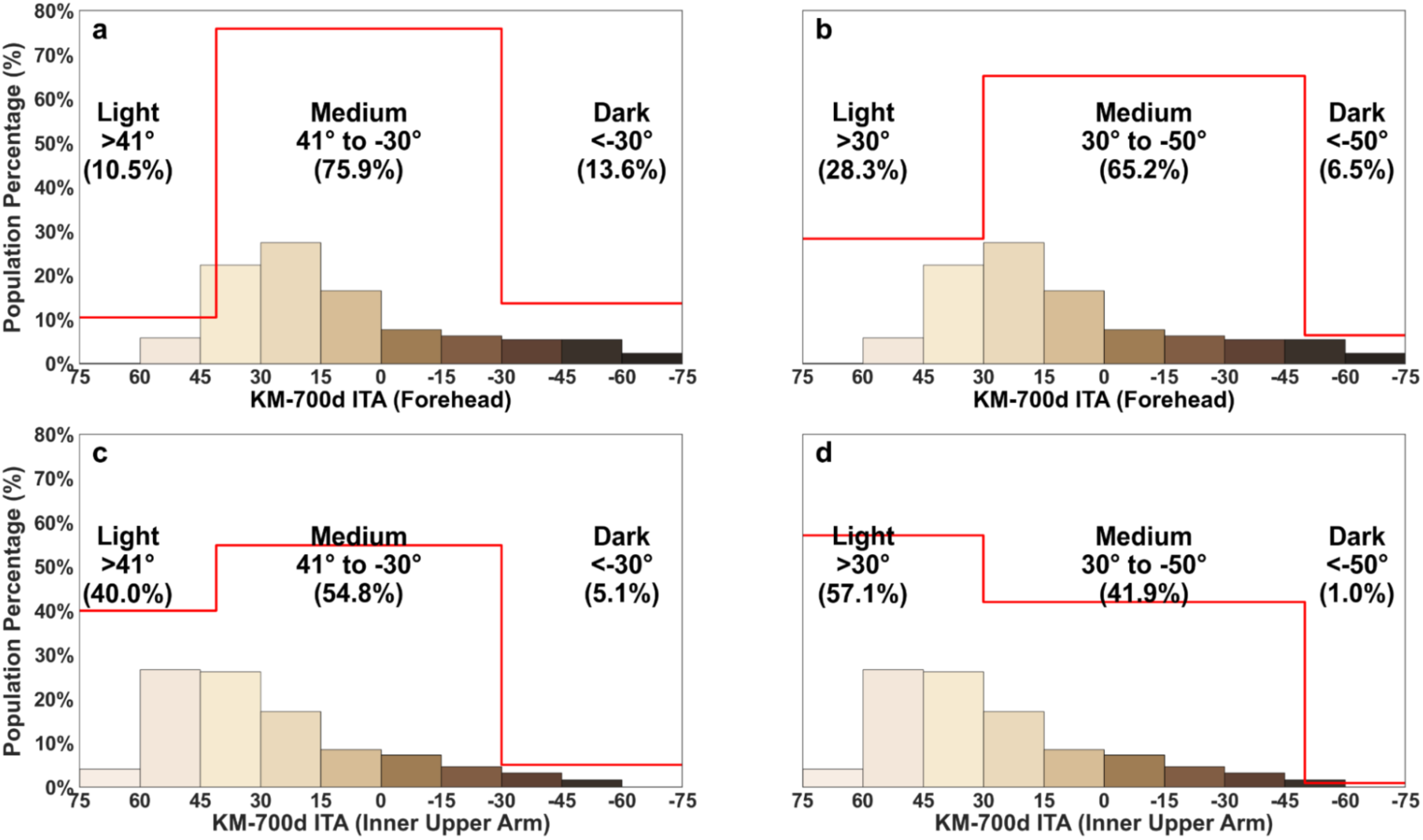
Study cohort diversity characterized by different individual typology angle cutoffs. This figure compares the proportion of the study cohort characterized as light, medium or dark when using different individual typology angle (ITA) thresholds. The x-axes show ITA measured by the Konica Minolta CM-700d spectrophotometer (KM-700d) at the defined anatomical site, and the y-axes show the population percentage within each category. Red stepped lines in each subplot outline the cumulative proportions of the cohort within the defined subgroups. ITA thresholds in a,c align with existing published thresholds for light and dark pigmentation, while thresholds in b,d are alternative thresholds.

#### Variability of pigmentation categorization (light, medium, or dark) by assessment site

The choice of anatomical site impacted the proportion of the cohort characterized as light, medium or dark (Figure 4b). Measurement at the dorsal DP (16.4% ITA<-30° and 9.1% <-50°) and forehead (13.6% ITA<-30° and 6.5% <-50°) resulted in a higher proportion characterized as dark, while the inner upper arm (5.1% ITA<-30° and 1.0% <-50°) and palmar DP (0.7% ITA<-30° and 0.1% <-50°) resulted in lower proportions (Figure S10).

Within any individual, ITA measured across anatomic sites demonstrated consistent trends and correlations (Table 3 and Figure 5). For example, sun-exposed sites tended to have lower ITA than sun-protected sites (ITA_forehead_=10.3°±30.3 < ITA_inner upper arm_ =27.4°±27.4, paired t-test *p*-value:<0.001), and ITA from sun-exposed sites showed closer absolute agreement than ITA between sun-exposed and sun-protected sites. For example, mean error and rho for ITA_dorsalDP_ vs ITA_forehead_ was 3.4° and 0.82, respectively, while for ITA_dorsalDP_ vs ITA_inner upper arm_ was -20.1° and 0.80, respectively. Sites with consistent repeat measures also correlated strongly (e.g., forehead and inner upper arm).

**Figure 5.**
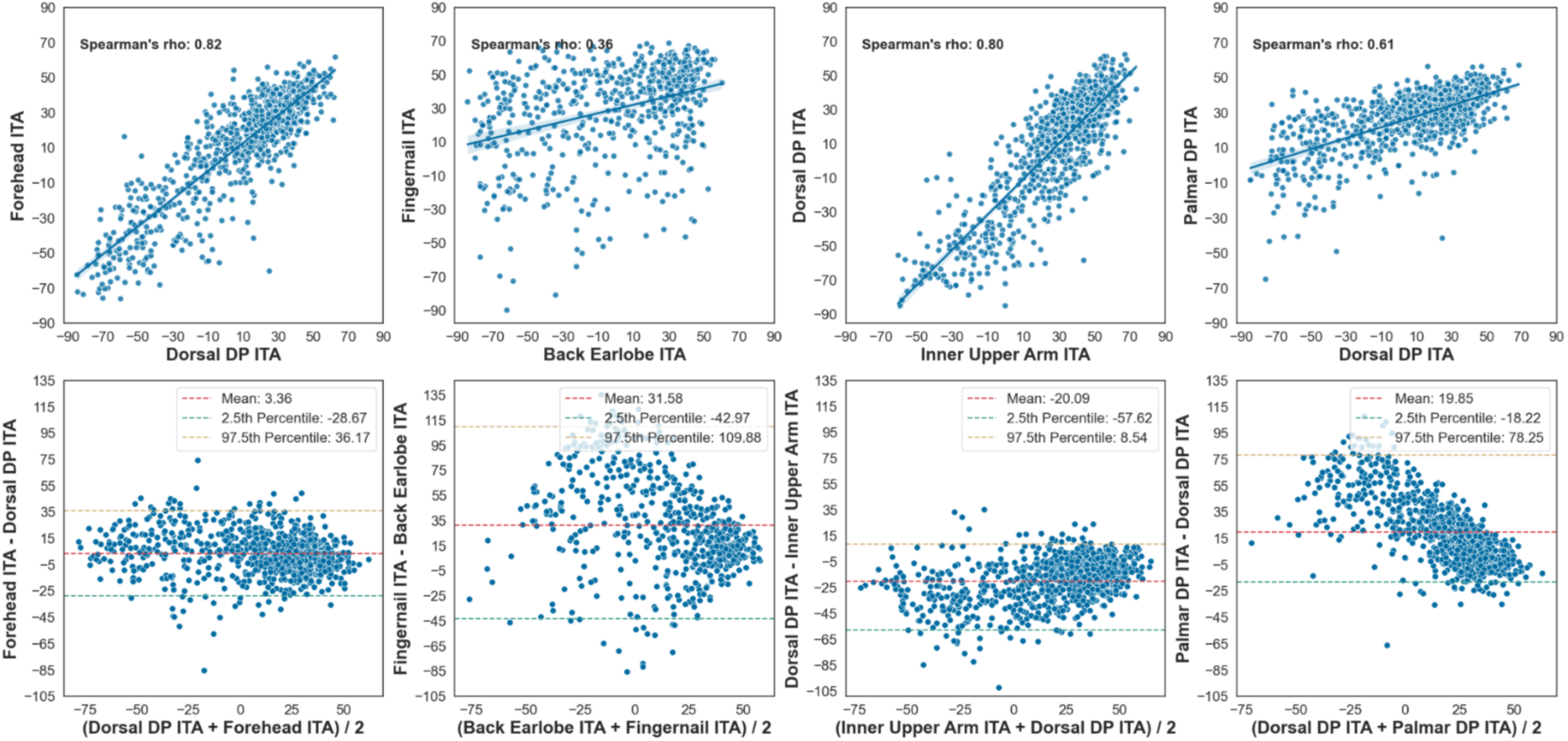
Comparison of individual typology angle between different anatomical sites. This figure consists of a four by two grid of subplots, with each column representing a pair of anatomical sites. The first column compares a pair of sites with high Spearman’s rho and a low mean difference of individual typology angle (ITA) measured by Konica Minolta CM-700d spectrophotometer (KM-700d). The second column shows the pair with the lowest Spearman’s rho. The third column compares a pair with high Spearman’s rho but a high mean difference of ITA. The fourth column represents the comparison between the palmar distal phalanx (DP) and dorsal DP. The top row displays regression plots, where the ITA from one site is plotted on the x-axis and the ITA from the paired site on the y-axis. Each regression plot includes a shaded area to visualize the 95% confidence interval for the estimated regression line and an annotation of Spearman’s rho for the correlation between the two sites. The bottom row contains Bland-Altman (BA) plots, with the x-axis representing the mean ITA of the two sites and the y-axis showing the difference in ITA between them. The BA plots are annotated with the mean difference and the 2.5th and 97.5th percentiles to illustrate the limits of agreement. Data shown are from both ICU and lab cohorts.

**Table 3.** Quantitative comparison of spectrophotometer individual typology angle between anatomical sites.

|  | Cheek | Dorsal DP | Fingernail | Forehead | Front Earlobe | Inner Upper Arm | Nare | Palmar DP |
| --- | --- | --- | --- | --- | --- | --- | --- | --- |
| Back Earlobe | 0.49<br>($<0.001$) | 0.52<br>($<0.001$) | 0.35<br>($<0.001$) | 0.51<br>($<0.001$) | 0.44<br>($<0.001$) | 0.51<br>($<0.001$) | 0.43<br>($<0.001$) | 0.4<br>($<0.001$) |
| Cheek | | 0.78<br>(0.11) | 0.45<br>($<0.001$) | 0.86<br>($<0.001$) | 0.68<br>(0.95) | 0.73<br>($<0.001$) | 0.76<br>($<0.001$) | 0.56<br>($<0.001$) |
| Dorsal DP | | | 0.43<br>($<0.001$) | 0.82<br>($<0.001$) | 0.7<br>(0.23) | 0.79<br>($<0.001$) | 0.71<br>($<0.001$) | 0.64<br>($<0.001$) |
| Fingernail | | | | 0.42<br>($<0.001$) | 0.37<br>($<0.001$) | 0.4<br>(0.02) | 0.41<br>($<0.001$) | 0.42<br>($<0.001$) |
| Forehead | | | | | 0.72<br>(0.02) | 0.83<br>($<0.001$) | 0.77<br>($<0.001$) | 0.56<br>($<0.001$) |
| Front Earlobe | | | | | | 0.64<br>($<0.001$) | 0.61<br>($<0.001$) | 0.48<br>($<0.001$) |
| Inner Upper Arm | | | | | | | 0.66<br>($<0.001$) | 0.53<br>(0.06) |
| Nare | | | | | | | | 0.51<br>($<0.001$) |

This table shows the strength of correlation and agreement of Konica Minolta CM-700d spectrophotometer (KM-700d) individual typology angle (ITA) measured at different anatomical sites for all study participants (ICU and lab cohorts) using the Spearman's rho ( $\rho$ ) and the paired t-test. Results are presented in the format of Spearman's $\rho$ (p-value from the paired t-test). Spearman's $\rho$ values $> 0.6$ are interpreted as strong correlations. Paired t-test p-values $< 0.05$ indicate that KM-700d ITA measurements at the two sites are significantly different, suggesting poor agreement.

#### Comparison of self-reported NIH race category with skin pigment measurement methods

For each self-reported race category (Table 1), we found a wide range of measured subjective scale categories and ITA values (Figure 6 and Table 4). While almost all MST HIJ subjects had self-identified as ‘Black or African American,’ we found that self-identified ‘Black or African American’ and self-identified ‘White’ participants have a wide range of MST values with significant overlap.

**Figure 6.**
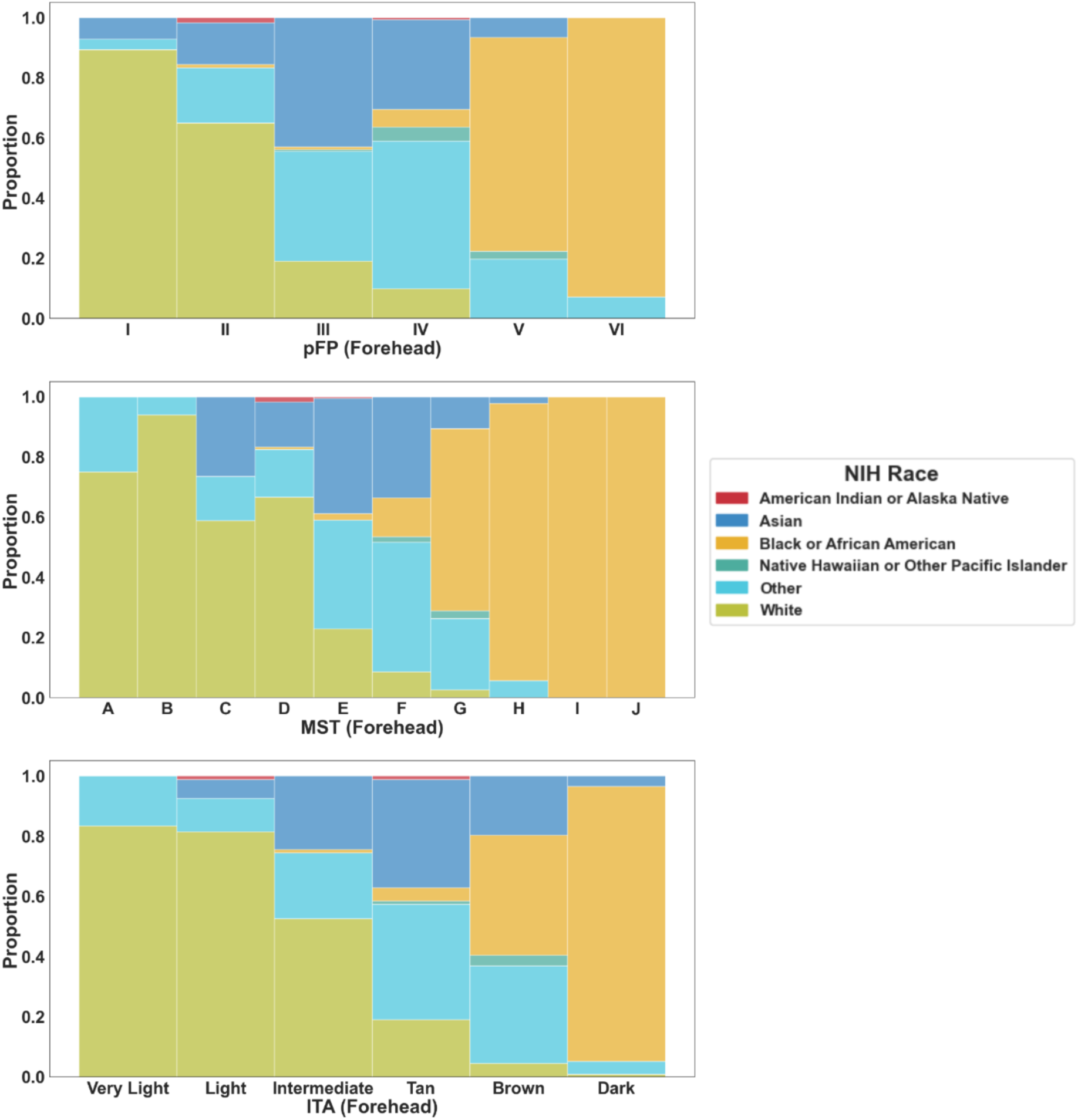
Distribution of pigmentation categorization by NIH race category. This figure includes three stacked histograms, each corresponding to one skin pigment assessment method (perceived Fitzpatrick Scale (pFP), Monk Skin Tone Scale (MST), and individual typology angle (ITA) measured by Konica Minolta CM-700d (KM-700d). Race categories are represented as uniquely colored segments within each bar. The x-axis shows the scale categories, and the y-axis indicates the proportion of each race within those categories in the combined ICU and lab cohort.

**Table 4.** Individual typology range by NIH race, pFP and MST categories.

|  | Category | KM-700d ITA<br>Median [Q1, Q3] | KM-700d ITA<br>Range |
| --- | --- | --- | --- |
| **NIH Race** | American Indian or Alaska Native | 15.67 [13.13, 24.38] | (12.53, 43.5) |
|  | Asian | 21.17 [10.75, 28.57] | (-54.37, 46.55) |
|  | Black or African American | -32.25 [-49.45, -15.99] | (-75.84, 38.79) |
|  | Native Hawaiian or Other Pacific Islander | 0.97 [-5.94, 9.15] | (-10.44, 20.61) |
|  | Other | 18.91 [6.77, 27.37] | (-62.1, 61.87) |
|  | White | 35.61 [27.96, 42.25] | (-32.41, 59.13) |
| **pFP**<br>**(Forehead)** | I | 41.48 [36.79, 49.27] | (21.06, 59.13) |
|  | II | 35.36 [29.08, 41.22] | (1.7, 61.87) |
|  | III | 24.6 [16.45, 30.19] | (-32.41, 49.38) |
|  | IV | 10.75 [3.18, 19.37] | (-51.95, 45.07) |
|  | V | -12.4 [-22.22, -2.37] | (-67.89, 21.31) |
|  | VI | -44.08 [-56.3, -29.2] | (-75.84, 21.49) |
| **MST**<br>**(Forehead)** | A | 50.08 [43.94, 56.15] | (38.03, 61.87) |
|  | B | 43.07 [38.79, 48.73] | (26.78, 56.61) |
|  | C | 41.29 [35.88, 47.81] | (6.77, 59.13) |
|  | D | 33.47 [27.67, 39.04] | (-32.41, 53.06) |
|  | E | 24.72 [17.15, 29.57] | (-10.68, 47.5) |
|  | F | 12.54 [4.42, 19.57] | (-26.35, 40.92) |
|  | G | -13.02 [-23.32, -5.61] | (-44.26, 12.87) |
|  | H | -44.49 [-54.51, -28.67] | (-70.25, -8.36) |
|  | I | -58.09 [-62.83, -42.46] | (-75.84, -25.73) |
|  | J | -72.61 [-72.96, -72.26] | (-73.31, -71.91) |

This table presents the median [Q1, Q3] and range of individual typology angle (ITA) values measured by Konica Minolta CM-700d (KM-700d) for each category of NIH race, perceived Fitzpatrick Scale (pFP), and Monk Skin Tone Scale (MST).

## Discussion

This study compares multiple skin pigmentation measurement methods and demonstrates that choice of method can cause significant variation in how the same cohort’s pigmentation is characterized, leading to potential underrepresentation of certain skin pigment populations. We found that ITA cutoffs commonly used to define ‘dark’ and ‘light’ pigmentation do not equitably represent real-world populations, and race and subjective scales have significant limitations that should restrict their use.

First we determined whether popular subjective methods can be used reliably to assess diversity of skin pigment. We found MST to have standardized colors with a wide range of luminosity, good discernibility across grouped pigmentation categories, and a reasonably consistent progression from light to dark that did not grossly under-represent certain pigment types. In contrast, VL and pFP had non-standardized colors, a more limited luminosity range, and the potential to under-represent darkly pigmented populations. Our findings on the limitations of subjective skin color scales align with prior studies.^19,20^

Second we determined whether different subjective and objective methods yield similar results when describing pigment of an individual or diversity of a cohort. We found significant differences when comparing ITA, VL, pFP, or MST, confirming prior reports that subjective scales can over- or under-represent darkly pigmented populations.^6^ When using an ITA<-30° to define dark, MST categories H-J most closely identified the proportion of the cohort with dark skin, with 77% of MST H-J participants having an ITA<-30° (Figure 7). The pFP overestimated the proportion of the cohort with dark skin, and only 51% of pFP V/VI participants had an ITA <-30°. While MST reasonably approximated a similar cohort proportion of dark participants as ITA (<-30°), two factors impact generalizability of this finding: 1. Operator bias is a known limitation of subjective scales and thus who the operator is (i.e., the person assigning the color category) can lead to over- or underrepresentation of pigment types;^21^ and, 2. We used existing ITA cutoffs that do not optimally characterize the range of human pigmentation.

**Figure 7.**
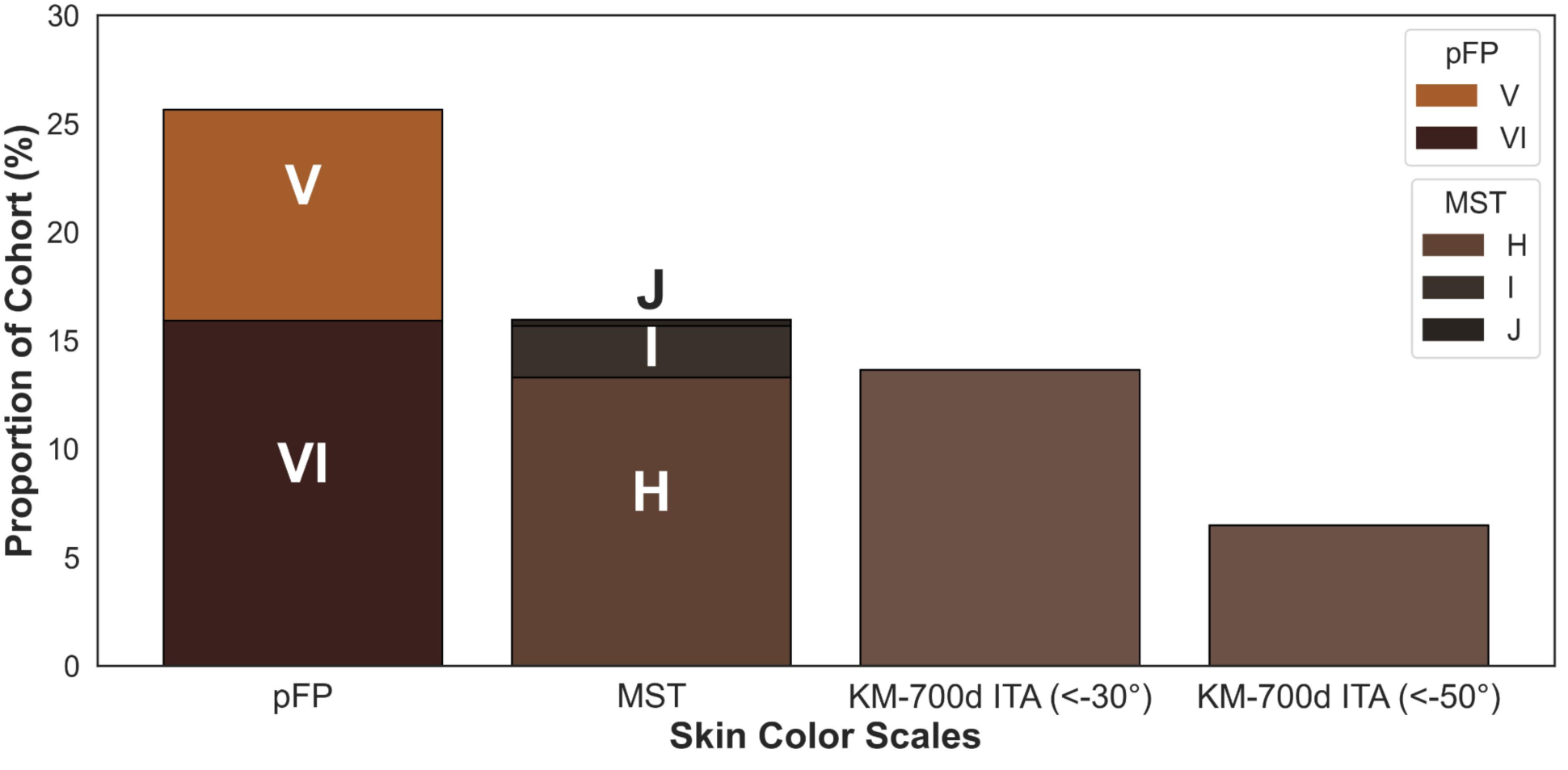
Proportion of cohort categorized as dark by method. The stacked histogram shows the proportion of the combined ICU and lab cohort classified as dark according to each method, with each segment representing a specific category (V and VI for pFP, and H, I, and J for MST). Each category is distinguished by a unique color for clarity.

Our results generated a new question: What ITA cutoffs equitably represent diverse skin pigments found worldwide? Existing ITA cutoffs are based on studies in sun-protected skin on the backs of mostly lightly pigmented volunteers and include six ITA categories ranging from very light (ITA>55°) to dark (ITA<-30°) based on the observation that the ITA categories roughly corresponded to FP phototypes I-VI, and measured ITA demonstrated a linear correlation with skin UV reactivity and melanin content.^15,16,22,23^ Given that this foundational work was conducted primarily in people with lighter skin, it stands to reason that current ITA pigmentation categories do not equitably represent the global population.

Given that our data are from a predominantly lightly-pigmented geography and still demonstrate a wide ITA range of -75.8° to 61.9°, with a significant proportion with ITA < -50°, we propose expanding existing ITA cutoffs to: very light ITA > 55°, light 55° ≥ ITA > 41°, intermediate light 41° ≥ ITA > 28°, tan 28° ≥ ITA > 10°, brown 10° ≥ ITA > -10°, intermediate dark -10° ≥ ITA > -30°, dark -30° ≥ ITA > -50°, and very dark -50° ≥ ITA (Figure 8). Adding two categories for dark pigment provides better parity with existing options to describe lighter pigment. Rather than split the colorspace into geometrically equal categories by ITA range, we retained categories for lighter populations to facilitate retrospective comparability and to acknowledge prior validation work that demonstrated correlation of melanin content to these categories.^15,24^ Based on our study’s geography, these cutoffs are likely conservative.

**Figure 8.**
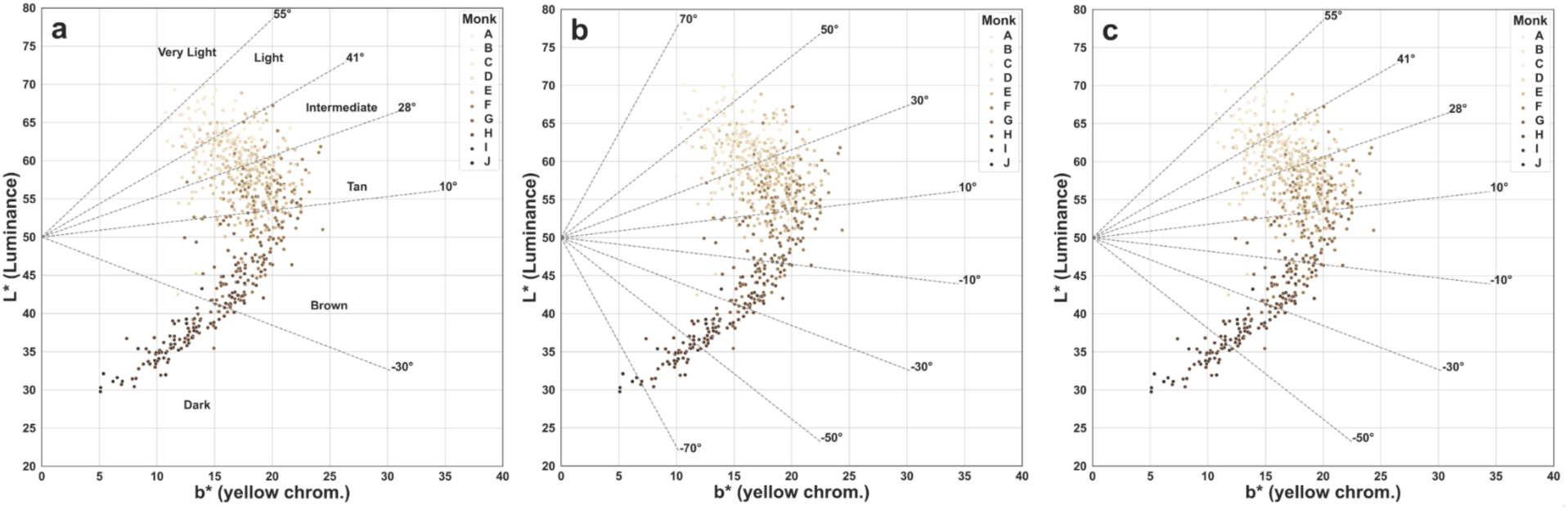
Proposed cutoffs for forehead L* vs b* for describing skin pigment. All panel display L* vs b* for all study participants (ICU and lab cohorts). Panel (**a**) uses previously published ITA cutoffs D<u>el Bino et al. (2013)</u>; panel uses geometrically equally divided bins, with each bin spanning 20°; and panel (**c**) Similar to (**a**), but with two additional bins with cutoffs at -10° and -50°.

Finally, our study provides data to guide selection of anatomic measurement sites. If it is not feasible to measure pigment at an anatomical site (e.g., a small finger or earlobe), then it is reasonable to use a surrogate site with similar sun exposure (e.g., forehead). The exceptions are the fingernail due to high SD in repeat measures, and the ventral hand (i.e., palm), due to poor absolute correlation of ITA with other sites. In our cohort, forehead assessment with MST and ITA was feasible and yielded reasonably reproducible data that correlated with common pulse oximetry measurement sites (i.e., finger, ear, nose).

This study has several limitations. First, data were collected in a predominantly lightly-pigmented geography and likely do not represent global populations. Additionally, we used self-identified NIH race categories and acknowledge that this social construct has many problems.^25^ Another limitation relates to data comparability because we used different protocols across study sites (e.g., anatomical sites, lighting, and number of raters) due to reliance on pre-existing studies. Furthermore, we did not assess for operator bias, a known limitation of subjective scales.^21^ We also used non-validated printing methods and two scales (pFP and VL) that lack standardized colors.^26^ Finally, we assessed performance of only two objective methods, and our finding of device comparability may not be widely applicable. Due to challenges with degradation of the DSCC white balance protective cap, the DSCC was used in relatively few participants. A problem during the study was found and rectified during interim analysis. Some repeat participants (mostly with darker pigmentation) in the lab cohort showed significantly different ITA on different days despite consistent MST. We traced the error to a discrete time period and a setting error on the KM-700d (MAV-SAV switch not matching the aperture). For all potentially affected participants, data were discarded and subjects were brought back for repeat measurements.

## Conclusions

Race should not be used to characterize skin pigment, and subjective scales should not be used alone to ensure diversity of skin pigment in research cohorts. Different methods applied to the same cohort yield different categorizations of light, medium, and dark, potentially overestimating diversity. The use of objective measures (e.g., ITA) with expanded categories to equitably characterize dark and light pigment ranges, is a pragmatic approach to ensuring diversity of skin pigment in research cohorts. Using a subjective scale with visually distinguishable bins spanning a wide range of standardized colors (e.g., MST) in addition to objective measures can help to more easily communicate findings and identify systematic problems with data collection (e.g., operator bias or spectrophotometer errors). Further work is needed to establish guidelines for equitable, objective assessments of skin pigmentation.

## Acknowledgements

We would additionally like to extend our sincere gratitude to Sandra Del Bino, Willem Verkruijsse, Jana Fernandez, Daryl Dorsey, Margaret Akey, Koyinsola Oyefeso, Alex Pogorzelski, Deleree Schornack, Shamsu Hashi, and Brandon Alford for their valuable contributions to this project.

## Data Availability Statement

All data supporting the findings of this study are openly accessible through the Open Oximetry Data Repository. Data are de-identified and shared under the repository’s terms, ensuring adherence to data collection protocols, local IRB approvals, and other relevant regulations. Users can access the data by creating a PhysioNet account and agreeing to the repository’s terms of use. For access details, please visit the Open Oximetry website.

## Ethics Statement

This study received approval from the University of California, San Francisco (UCSF) Committee on Human Research IRB, with protocols #21-35637, #22-36553, and #23-40212. Written informed consent was obtained from all participants following the Declaration of Helsinki guidelines. Data from the Open Oximetry Data Repository were fully de-identified to protect participant privacy.

## Contributor Statement

KM, LS, JF, ML, EM, TL, PB, EI, RB, OO, FN contributed to conception and design of the work. GL, EB, SE, IA, SH, YC, CH, ML, LO contributed to data acquisition. ML, LS, TL, DC, JF, EM contributed to data analysis. ML, LS, LO, FN led drafting of the manuscript, and all authors participated in data interpretation and critically reviewing the work and approving the final version.

- What is already known about this topic?

- Multiple methods exist for characterizing human skin pigment, but there is no consensus or standardized approach, and existing scales are widely known to be misused. Recently, the lack of diversity in medical device regulatory validation testing cohorts has been scrutinized as potentially contributing to pulse oximeters not working as well in people with dark skin pigment.
- What does this study add?

- This study demonstrates the strengths and limitations of popular subjective and objective skin pigmentation measurement methods, including the potential for some methods to falsely overstate the diversity of study cohorts. The study also proposes new ITA cutoffs that better reflect real world populations and better represent people with dark skin pigment.
- What is the translational message?

- The study’s findings support changes in common approaches to characterizing human skin pigment for the purpose of characterizing study cohorts. If integrated into regulatory requirements and applied to medical device research cohorts, these improved approaches could result in more equitable device design and less health disparities.

---

**Figure S1.**
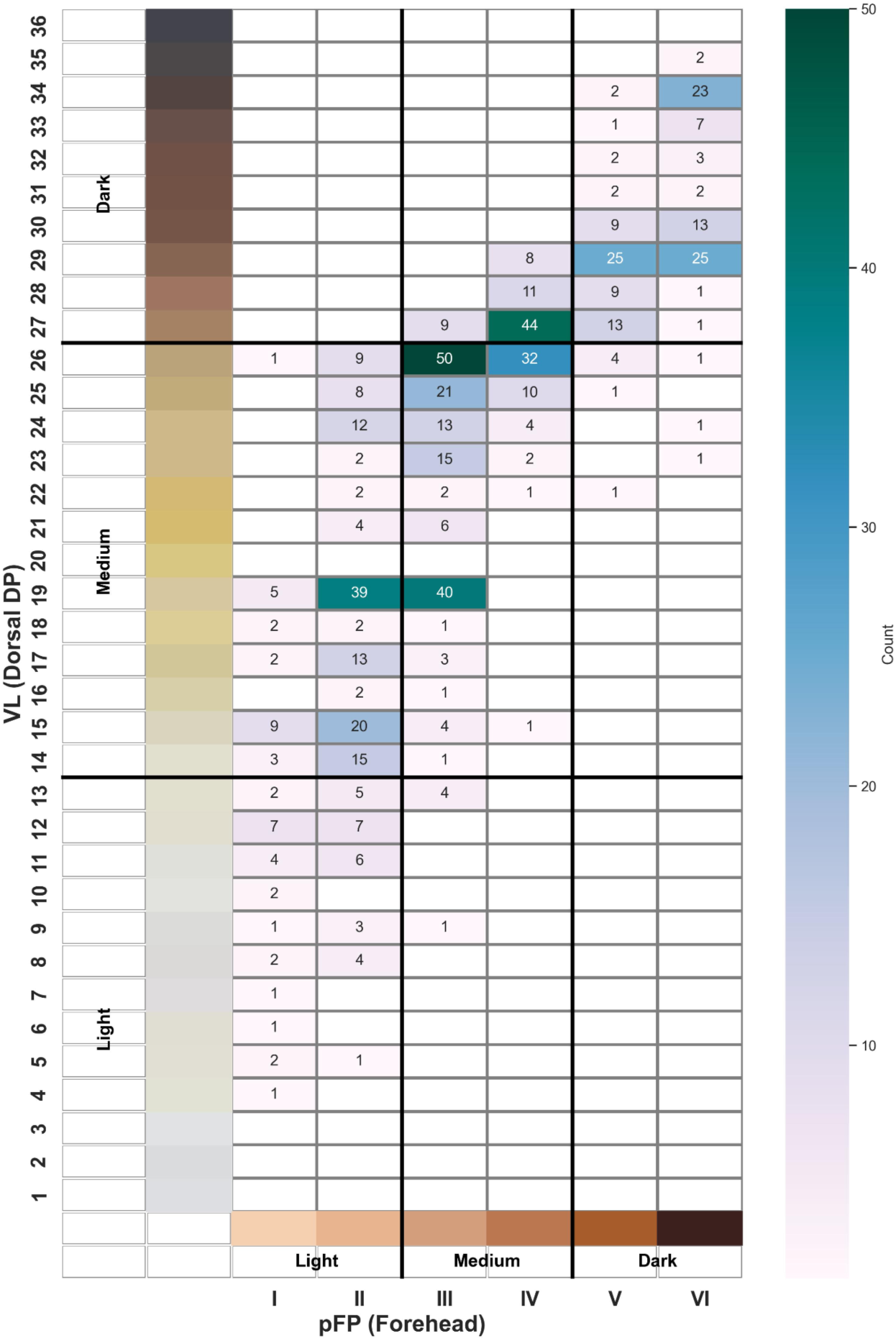
Comparison of participant categorization by perceived Fitzpatrick Scale vs. Von Luschan Scale. The heatmap shows the count of subjects in the combined ICU and lab cohort for matched perceived Fitzpatrick Scale (pFP) and Von Luschan Scale (VL) categories, with darker green colors representing higher counts. The pFP scale was assessed at the forehead, while the VL scale was assessed at the dorsal distal phalanx (DP). Bold lines divide each scale into three categories: light, medium, and dark, to help visualize the alignment of these categories defined by each scale. Data shown are from ICU cohort only, as the VL scale was not used in the lab cohort. Many participants were categorized differently depending on which scale was utilized.

**Figure S2.**
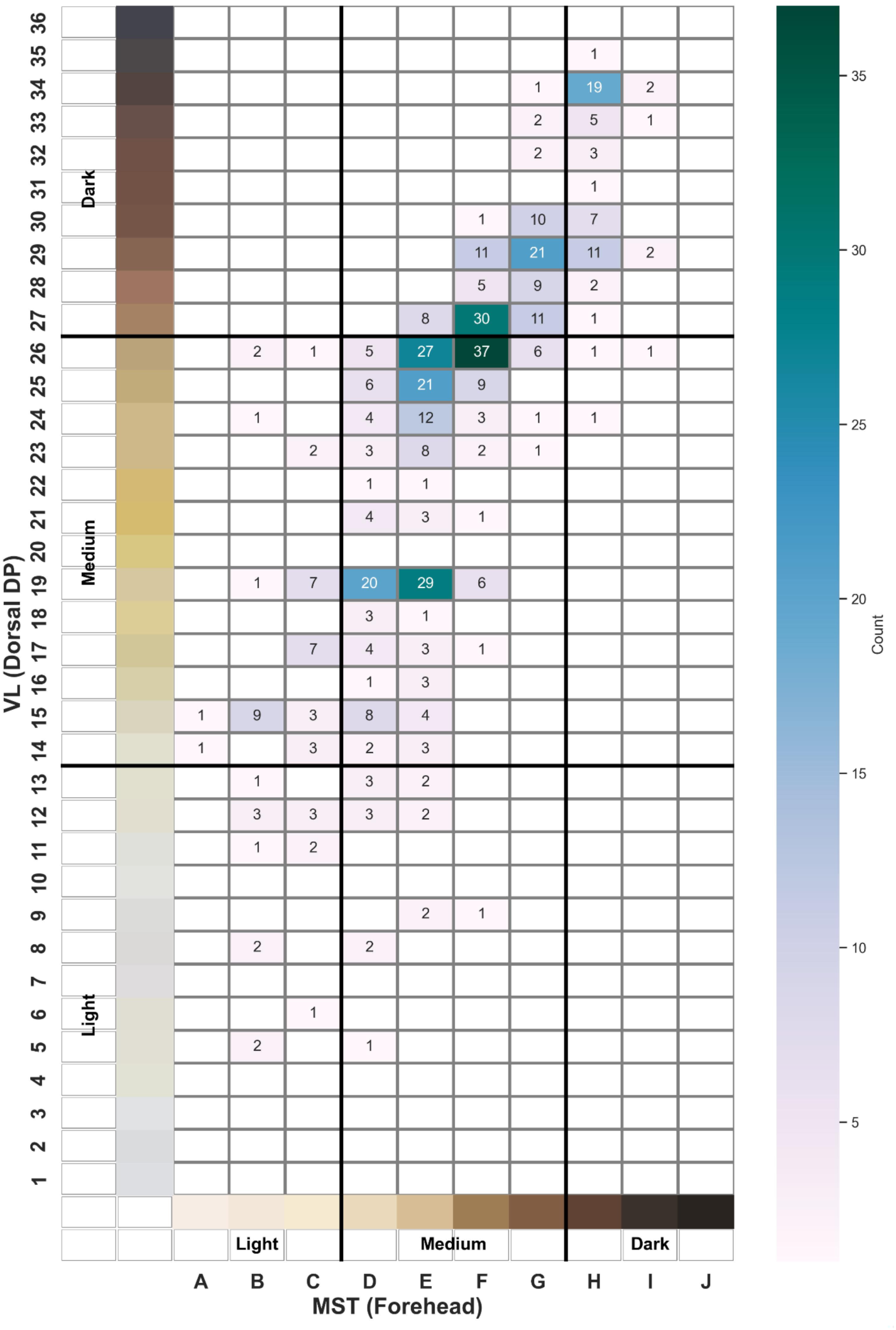
Comparison of participant categorization by Monk Skin Tone Scale vs. Von Luschan Scale. The heatmap shows the count of subjects in the combined ICU and lab cohort for matched Monk Skin Tone Scale (MST) and Von Luschan Scale (VL) categories, with darker green colors representing higher counts. The MST scale was assessed at the forehead, while the VL scale was assessed at the dorsal distal phalanx (DP). Bold lines divide each scale into three categories: light, medium, and dark, to help visualize the alignment of these categories defined by each scale. Data shown are from ICU cohort only, as the VL scale was not used in the lab cohort. Many participants were categorized differently depending on which scale was utilized.

**Figure S3.**
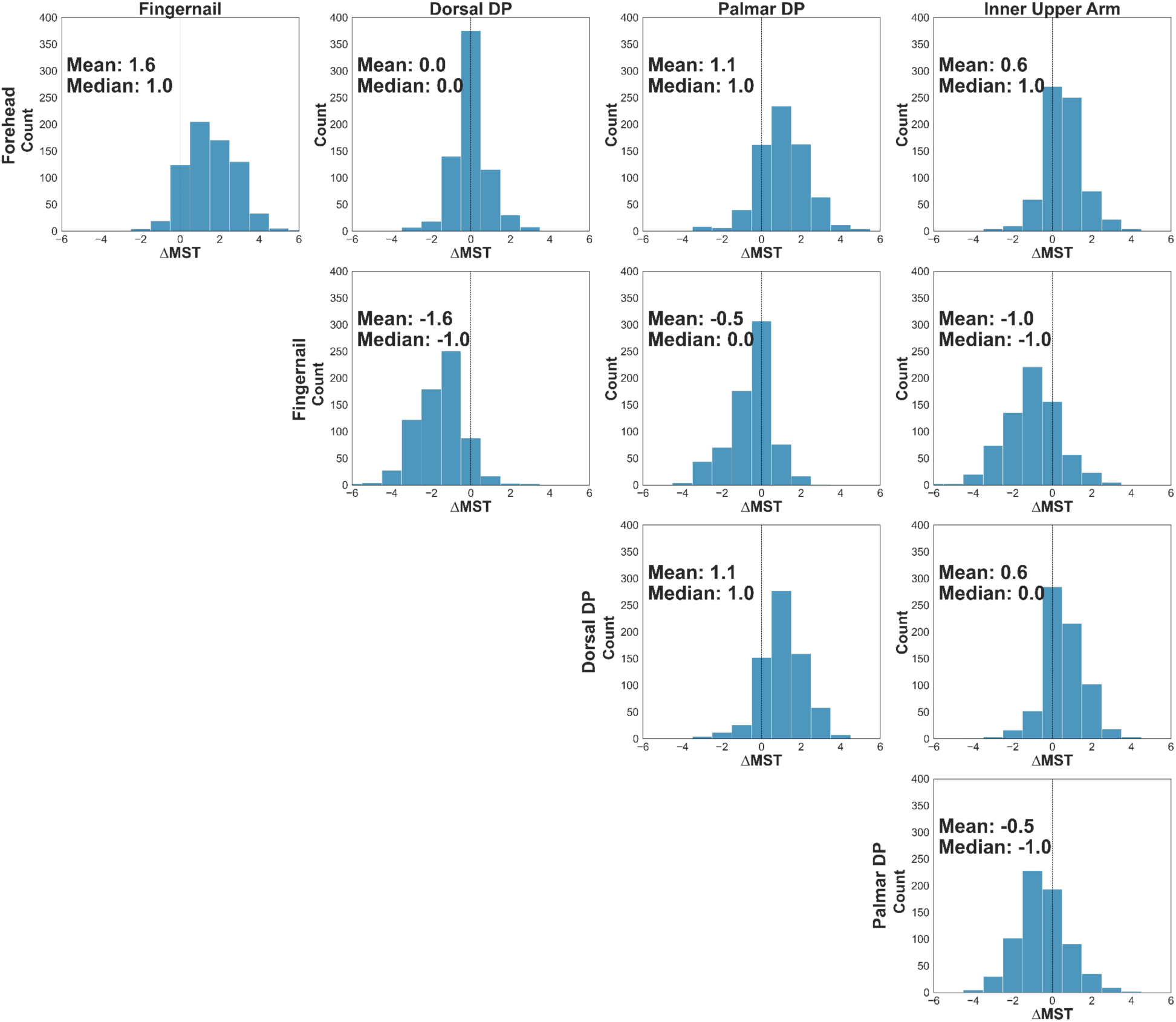
Comparison of the difference in MST (ΔMST) measured at separate anatomic sites. These histograms visualize assigned ΔMST across different anatomical measurement sites (indicated by the row and column labels) within the same subject across the entire subject cohort. For each anatomic site comparison, the histogram provides a count of the number of subjects with a given ΔMST between anatomic sites, and the mean median ΔMST is shown. Closest agreement in MST between sites occurs between forehead and dorsal DP (ΔMST centered at 0, mean and median MST= 0).

**Figure S4.**
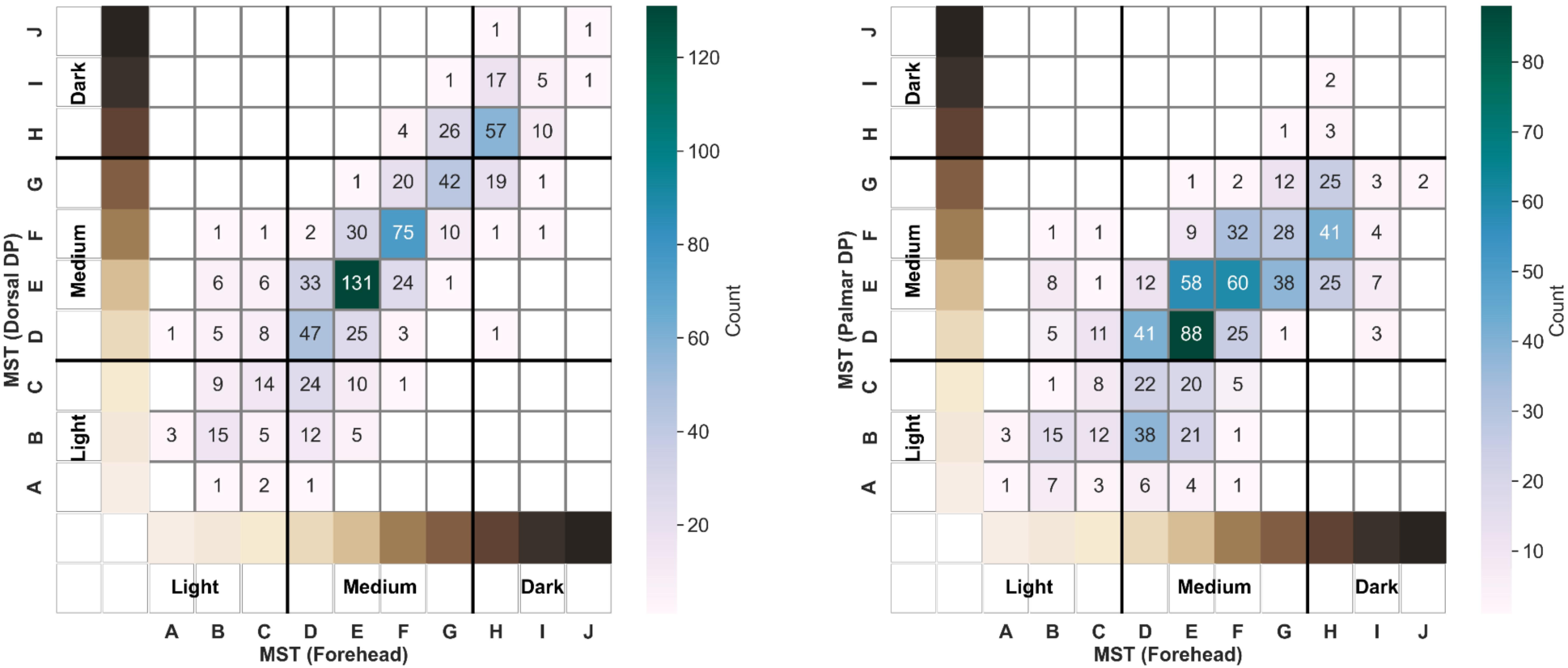
Comparison of participant categorization by Monk Skin Tone Scale across different anatomical sites. This figure includes two heatmaps showing the count of participants in the combined ICU and lab cohorts for matched Monk Skin Tone Scale (MST) categories assessed at different anatomical sites, with darker green colors representing higher counts. The first heatmap compares MST assessed at the forehead and dorsal distal phalanx (DP), both representing sun-exposed sites. The second heatmap compares MST measured at the forehead (a sun-exposed site) and palmar DP (a sun-unexposed site).

**Figure S5.**
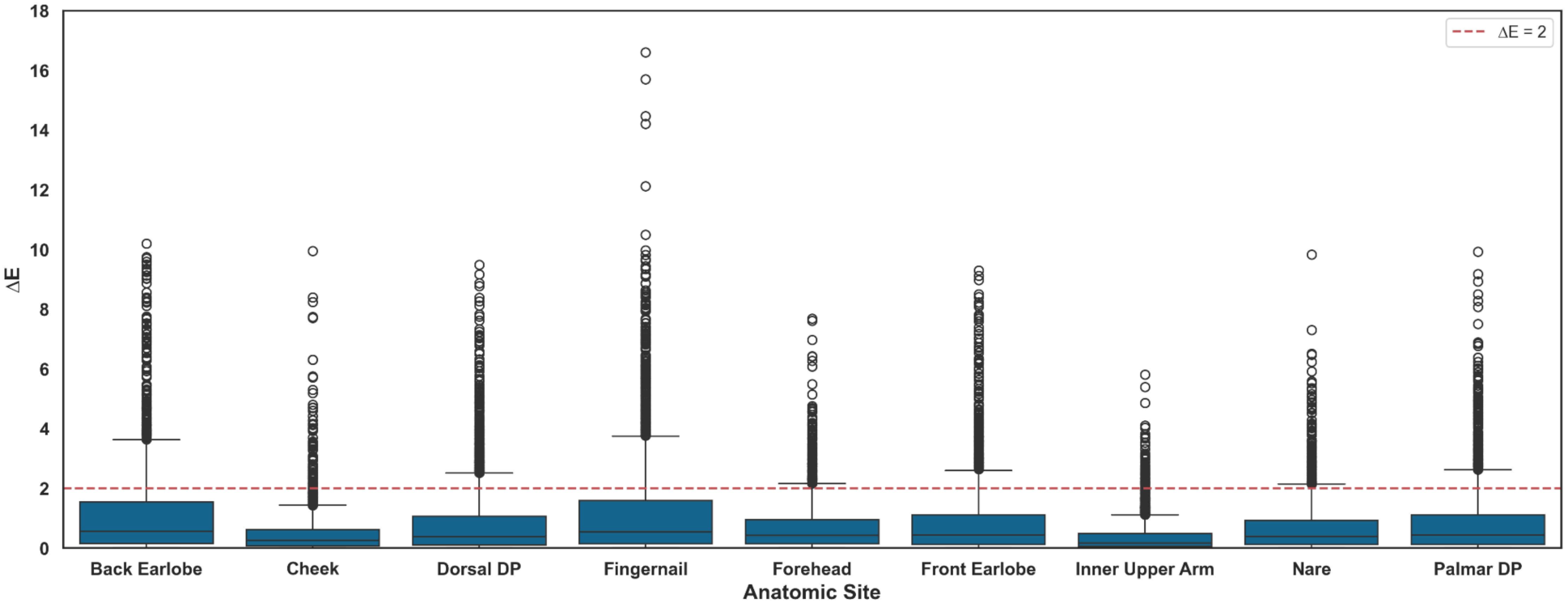
Distribution of ΔE among triplicate L*a*b* measures for each anatomical site. This figure presents boxplots showing the distribution of ΔE values among triplicate L*a*b* measures from the Konica Minolta CM-700d (KM-700d) for each anatomical site in the combined ICU and lab cohort. Each boxplot represents a specific anatomical site. The red dashed line indicates a ΔE of 2, a commonly used threshold for perceptibility of color difference.

**Figure S6.**
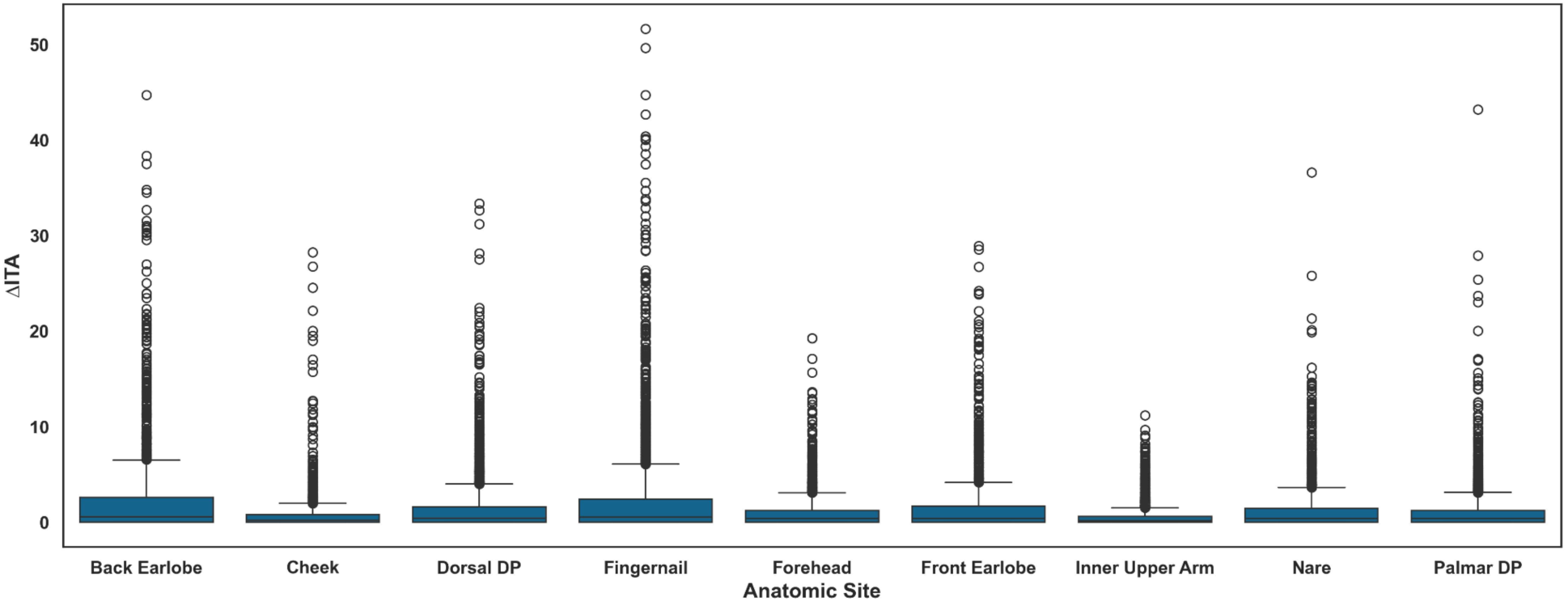
Distribution of ΔITA among triplicate ITA measures for each anatomical site. This figure presents boxplots showing the distribution of ΔITA values among ITA triplicates from the Konica Minolta CM-700d (KM-700d) for each anatomical site in the combined ICU and lab cohort. Each boxplot represents a specific anatomical site. Sites that are more challenging to measure (fingernail, back of earlobe) exhibit more significant outliers than other sites.

**Figure S7.**
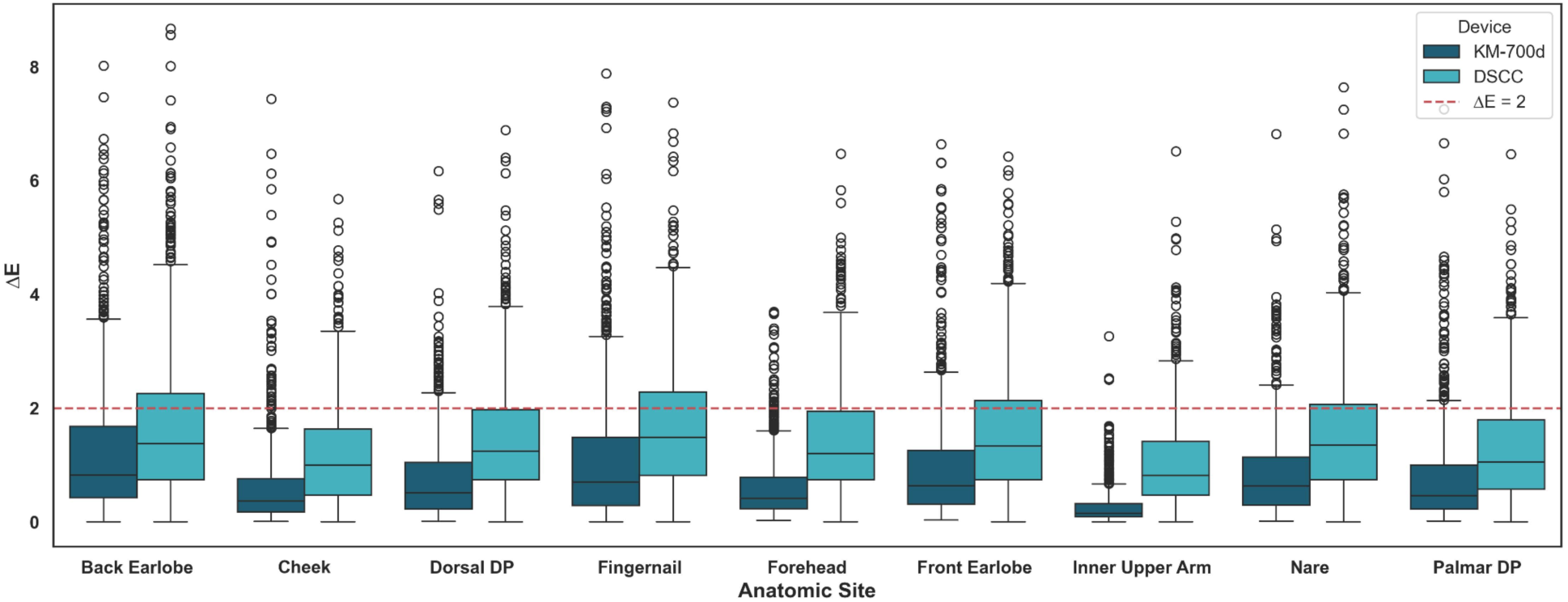
Comparison of ΔE among triplicate L*a*b* between two objective methods across anatomical sites. This figure presents boxplots showing the distribution of ΔE values among L*a*b* triplicates for each anatomical site, each objective method used (Konica Minolta CM-700d spectrophotometer (KM-700d), and Delfin Skin Color Catch colorimeter (DSCC). Each anatomical site is represented by two boxplots: one for the KM-700d device (darker blue) and one for the DSCC device (lighter blue). The red dashed line indicates a ΔE of 2, a commonly used threshold for perceptibility of color difference. Data shown are from ICU cohort only, as the DSCC device was not used in the lab cohort.

**Figure S8.**
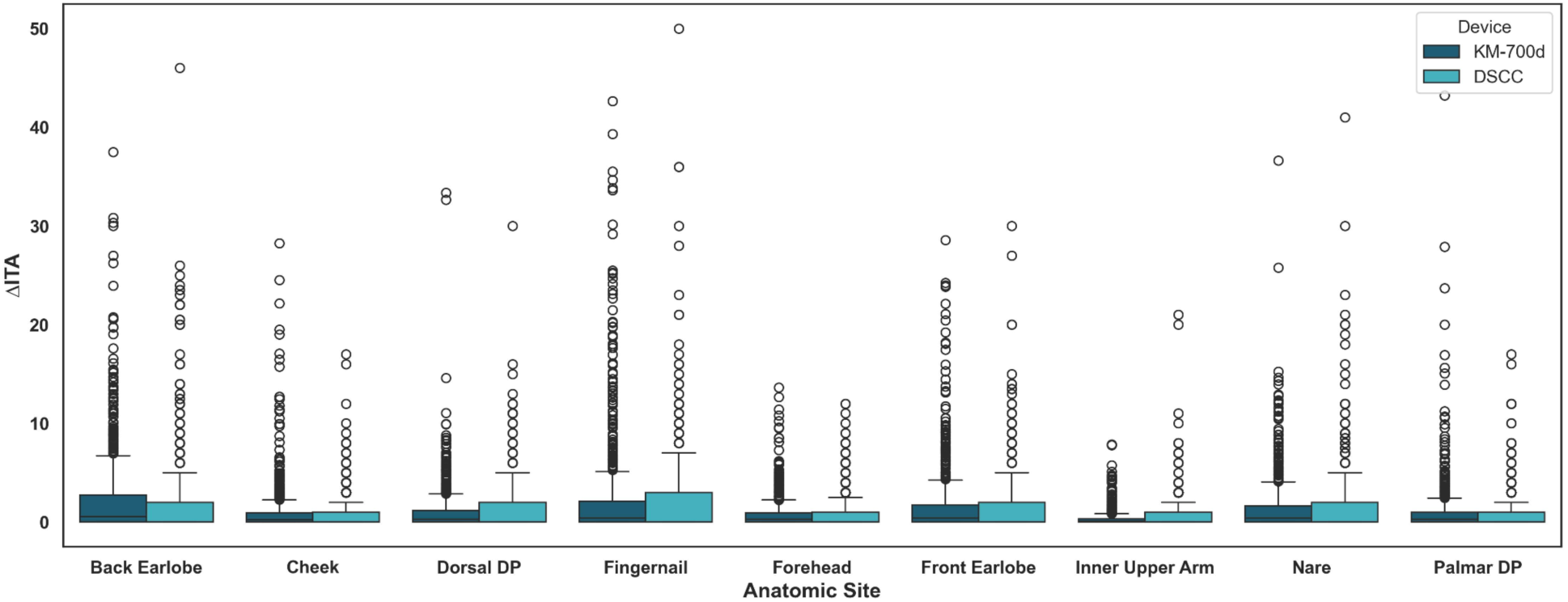
Comparison of ΔITA among ITA triplicates between two objective methods across anatomical sites. This figure presents boxplots showing the distribution of ΔITA values among ITA triplicates for each anatomical site, each objective method used (Konica Minolta CM-700d spectrophotometer (KM-700d), and Delfin Skin Color Catch colorimeter (DSCC)). Each anatomical site is represented by two boxplots: one for the KM-700d device (darker blue) and one for the DSCC device (lighter blue). Data shown are from ICU cohort only, as the DSCC device was not used in the lab cohort.

**Figure S9.**
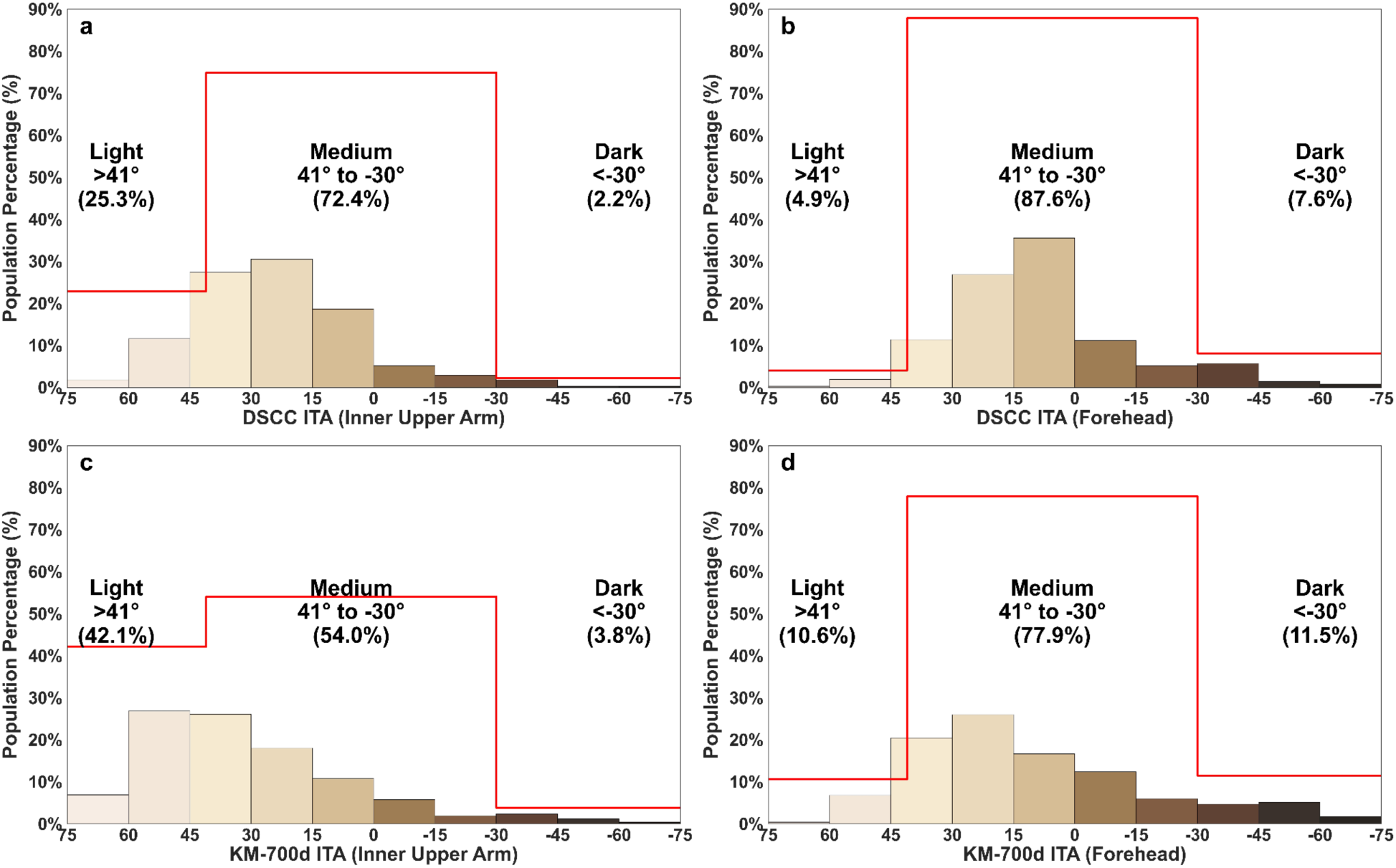
Distribution of ITA in different anatomical sites by device. This figure compares the proportion of the study cohort characterized as light, medium or dark when measured by different colorimeters. The x-axes shows ITA measured by Delfin Skin Color Catch colorimeter on the top row and measurement by Konica Minolta CM-700d spectrophotometer (KM-700d) on the bottom row, and the y-axes show the population percentage within each category. Red stepped lines in each subplot outline the cumulative proportions of the cohort within the defined subgroups. ITA thresholds in a,c align with existing published thresholds for light and dark pigmentation, while thresholds in b,d are alternative thresholds.

**Figure S10.**
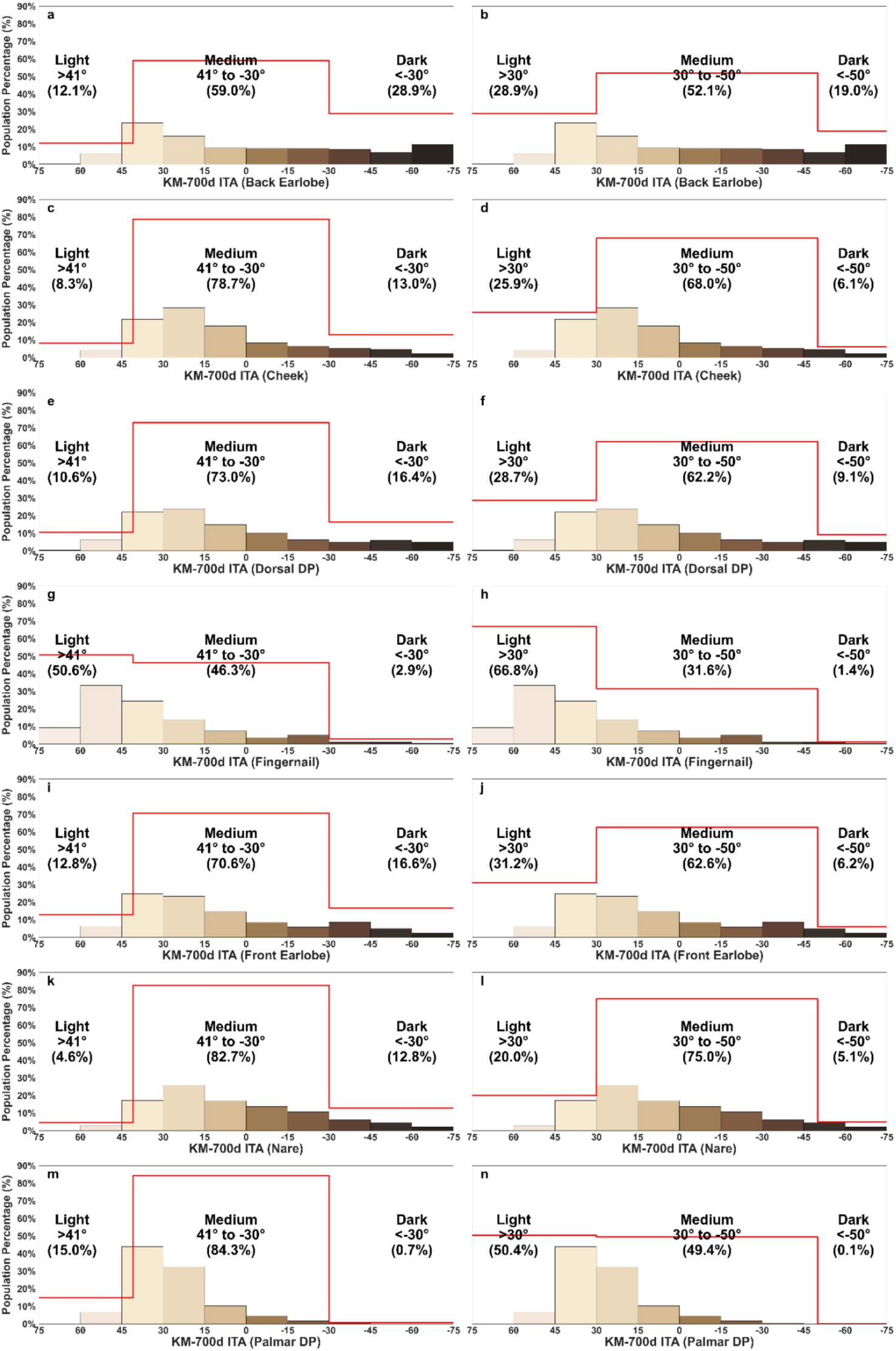
Distribution of KM ITA in different anatomical sites. This figure compares the proportion of the study cohort characterized as light, medium or dark when using different individual typology angle (ITA) thresholds. The x-axes show ITA measured by the Konica Minolta CM-700d spectrophotometer (KM-700d) at the defined anatomical site, and the y-axes show the population percentage within each category. Red stepped lines in each subplot outline the cumulative proportions of the cohort within the defined subgroups. ITA thresholds in a, c, e, g, i, k, m align with existing published thresholds for light and dark pigmentation, while thresholds in b, d, f, h, j, l, n are alternative thresholds.

